# Multidimensional cognitive deficits in the typical and atypical variants of Alzheimer’s disease

**DOI:** 10.1101/2025.03.04.25323342

**Authors:** Shalom K. Henderson, Alexander G. Murley, Thomas E. Cope, Lucy Bowns, Maura Malpetti, Karalyn E. Patterson, James B. Rowe, Matthew A. Lambon Ralph

## Abstract

In this two-part investigation, we examined whether Alzheimer’s disease (AD) phenotypes are distinct clinical entities or represent positions within a graded multidimensional space. First, using a large retrospective dataset of past research participants (*n* = 413) from memory clinics, we examined the comparative distributions of cognitive performance in people diagnosed with typical amnestic AD (tAD), logopenic variant of primary progressive aphasia (lvPPA), and posterior cortical atrophy (PCA). Secondly, a prospective deep phenotyping study of lvPPA (*n* = 18) compared to typical AD (*n* = 9) addressed the following questions: (1) Does the multidimensional cognitive pattern of impairment only emerge in advanced lvPPA, and how does it compare to tAD? (2) Do memory deficits in lvPPA appear in a simple clinic-level cognitive assessment or require in-depth neuropsychological investigation? (3) To what extent is performance on verbal episodic memory attributable to language impairment? (4) Do the patterns of decline in lvPPA and tAD stay categorical or multidimensional over time? We explored the associations between scores derived from a principal component analysis of cognitive measures, and grey matter volumes in key memory- and language-related brain regions, at baseline and longitudinally. The clinic-level assessment revealed similar results in both the prospective and retrospective data: (i) patients showed graded distinctions (e.g., predominant visual versus language impairment in people with PCA versus lvPPA) and overlap (e.g., shared weakness in domains such as memory); and (ii) people with lvPPA and tAD were equally impaired on <u>both</u> verbal and non-verbal memory tests in lvPPA and tAD. Longitudinal assessment showed phenotypic dispersion: (i) people with tAD showed varied patterns of phenotypic differentiation; and (ii) people with lvPPA and lvPPA+ exhibited a multidimensional pattern of decline with decreasing principal component scores and worsening multi-domain cognitive performance. The results of Bayesian linear regressions showed evidence for the association of grey matter volumes in language and memory networks, including the bilateral hippocampi, precuneus, posterior cingulate, and temporo-parietal regions with principal component analysis derived scores. The graded distinctions amongst typical amnestic and atypical (language and visual) phenotypes of AD support the proposal for a transdiagnostic, multidimensional phenotype geometry that spans all AD subtypes. We argue that it is essential to include all AD phenotypes in clinical trials and treatments, with a particular focus on transdiagnostic symptoms, considering their relevance to disease burden and interventions.

## Introduction

The phenotypes of Alzheimer’s disease (AD) extend beyond amnesia and may include dominant difficulties in language, visual, executive, behavioural and motor domains.^1–4^ There is ongoing debate about how to consider these AD phenotypes. A classical diagnostic perspective is based on categorical boundaries.^5–8^ However, recent studies have increasingly reported graded variations within and between the AD phenotypes, leading to the hypothesis that AD subtypes represent positions within a graded multidimensional space.^3,9–11^ It is difficult to draw definitive conclusions between these two hypotheses because there are confounding factors that include, but are not limited to, the severity of patients, granularity of testing, nature of the assessment, and differences in study design (e.g., cross-sectional versus longitudinal).

The logopenic variant of primary progressive aphasia (lvPPA) is almost invariably caused by AD pathology and is therefore also known as the “language-variant AD”. People with lvPPA present with impaired sentence repetition and single-word retrieval, as well as frequent phonological errors, and the condition is typically associated with left-predominant temporoparietal atrophy.^12–14^ Increasing disease severity is often regarded as the cause of multi-domain cognitive deficits in lvPPA.^15^ Episodic memory impairment, in particular, is reported to arise later in the condition.^16^ However, multiple targeted studies have found that even mild patients meeting diagnostic criteria for lvPPA exhibit episodic memory deficits when assessed with detailed neuropsychological testing and/or in-depth carer interviews.^17,18^ These discrepancies make it challenging to ascertain whether the multimodal pattern of impairment is present only in advanced cases or also in mild lvPPA, and how it relates to typical AD. There are also potential issues with the granularity of testing. In other words, can one distinguish a selective *versus* multidimensional pattern of impairment in a clinic-level assessment, or is an extended neuropsychological battery required to reveal the cognitive dimensions? Furthermore, the presence of true memory deficits in lvPPA has been disputed in the literature due to the potential contribution of aphasia to reduced verbal episodic memory test performance.^14,19^ If impaired memory performance relates to the verbal nature of the test materials, then individuals with lvPPA would be expected to perform significantly better on non-verbal than verbal episodic memory tests (relative to people with tAD). This hypothesis has not been formally tested in the current literature. Lastly, the majority of lvPPA research has been cross-sectional. Therefore, whether people with lvPPA remain categorically distinct, or progress in a multidimensional trajectory of decline, remains elusive given the paucity of longitudinal studies and their mixed findings.

Posterior cortical atrophy (PCA) is another well-characterized phenotypic variant of AD (sometimes referred to as the “visual-variant AD”) with spatial and object perception deficits, constructional apraxia, environmental agnosia, alexia, as well as features associated with Balint and Gerstmann syndromes.^6,20^ Similar to lvPPA, additional cognitive impairments are often reported in PCA, even in those who are mild, and the pattern of cognitive impairments mirrors that of tAD with impairments across the language, executive function, and memory domains.^9,20,21^ There is overlap in language profiles between PCA and lvPPA patients including word retrieval and verbal working memory difficulties.^22,23^ The fact that PCA and lvPPA (i) are the two most prominent and well-described, yet apparently phenotypically-contrastive, subtypes of AD^24^ and (ii) are associated with posterior atrophy (temporo-parietal in lvPPA and parieto-occipital in PCA)^25^ motivated our study to directly compare the cognitive performance of patients with PCA, lvPPA, and typical AD, which is rarely done in the literature.

In our two-part investigation, we first examined the comparative distributions of cognitive performance in people diagnosed with typical amnestic AD, lvPPA, and PCA using a large retrospective dataset of past research participants (*n* = 413) from the memory clinics of the Cambridge University Hospitals NHS Trust. We focussed on the particular domain of impairment that is associated with each AD phenotype (memory in typical AD, language in lvPPA, and visual in PCA) and asked whether the multidimensional pattern of cognitive impairment is present across the three AD phenotypes. Second, we conducted a prospective deep phenotype study of lvPPA compared to typical AD, which is an important contrast that acts as a test case of the broader issue of how to understand the relationship between atypical AD phenotypes and classical amnesia-led AD. We addressed the following questions: (1) Is the multidimensional cognitive pattern of impairment only present in advanced lvPPA, and how does it compare to tAD? (2) Do memory deficits in lvPPA appear in a simple clinic-level cognitive assessment or require in depth research-level investigation? (3) To what extent is performance on verbal episodic memory attributable to language impairment? (4) Do the patterns of decline in lvPPA and typical AD stay categorical or multidimensional over time? In addition, we explored the associations between principal component analysis derived scores and grey matter volumes in key memory- and language-related brain regions, at baseline and longitudinally. If typical and atypical AD phenotypes show graded overlap across different cognitive domains, this would offer valuable insights about the multidimensional nature of cognitive impairments resulting from AD.

## Materials and methods

### Part 1. Retrospective study of lvPPA, PCA and tAD

#### Participants

Our retrospective data included research participants (*n* = 413) with a clinical diagnosis of tAD,^26^ lvPPA,^12,27^ and PCA^6^ at memory clinics of the Cambridge University Hospitals NHS Trust who completed the Addenbrooke’s Cognitive Examination – Revised (ACE-R).

#### Assessments

The ACE-R was administered as a clinic-level cognitive assessment. In addition to the total score (max=100), the ACE-R is made up of domains of higher cognition with sub-scores in attention (/18), memory (/26), fluency (/14), language (/26), and visuospatial (/16).^28^

#### Statistical analysis

To test for group differences in age and sex, we conducted a Bayesian ANOVA followed by a *post hoc t*-test for pairwise multiple comparisons. We analysed the ACE-R data in two ways. First, as a proxy for overall severity, we stratified each diagnostic group into ranges based on ACE-R total scores. The four ranges were based on published cut-off values for the ACE-R: (1) upper limit of normal range (i.e., within 1 standard deviation of the control mean), (2) lower limit of normal range (i.e., between 1 and 2 standard deviations of the control mean), (3) below the cut-off (i.e., 3 standard deviations below the control mean), and (4) significantly below the cut-off (i.e., more than 3 standard deviations below the control mean). Using these four ranges, we tabulated the proportion of patients who were within normal range and below the cut-off for each of the ACE-R cognitive domains. This allowed us to examine how many people within each diagnostic group, as well as within each level of severity (as indexed by overall performance), had scores below the cut-off for all cognitive domains. Next, we conducted a Bayesian ANOVA to test for group differences in the total ACE-R scores, as well as in each cognitive ACE-R sub-domain, followed by post *hoc* independent samples *t*-test.

### Part 2. Prospective study of lvPPA and tAD

#### Participants

As described in Henderson *et al*.,^29^ twenty-seven people diagnosed with lvPPA (*n* = 18) and amnestic-led AD (*n* = 9), and twelve age-matched healthy controls participated in the study. Patient participants were recruited from specialist clinics for memory and language disorders at Cambridge University Hospitals NHS Trust. All participants were self-reported “White” and reported English as their first language (in keeping with East of England demography of older adults). Nine patients had tAD,^26^ ten patients met strict criteria for PPA and lvPPA,^12,27^ and eight patients were classified as “lvPPA+” as they previously satisfied definitions of PPA and lvPPA but by the time of this study had developed multi-domain cognitive impairments. All participants gave written informed consent in accordance with the Declaration of Helsinki, with patient participants supported by family or caregiver when necessary. Biomarker evidence of Alzheimer-type was available for a subset of participants. Four patients (1 tAD, 1 lvPPA, 2 lvPPA+) had AD confirmatory status with plasma pTau217 > 0.63 pg/mL^30^ and another lvPPA patient had cerebrospinal fluid biomarkers indicative of AD.

At the initial assessment, one lvPPA+ patient did not complete the ACE-R due to difficulty following task instructions. In addition, specific episodic memory tests, namely the Rey Auditory Verbal Learning Test (RAVLT) and Rey Osterrieth Complex Figure (ROCF), were attempted but discontinued in five and two patient participants, respectively, due to task difficulty. Details about missing data are included in the statistical analysis section. Nine patient participants (tAD *n* = 3; lvPPA *n* = 2; lvPPA+ *n* = 4) were followed longitudinally. Cognitive assessment using the same test battery was attempted on two separate occasions. Two patients were excluded from the longitudinal behavioural analysis as one patient was unable to complete any of the cognitive tests and another patient only completed a confrontation naming test. The mean time between initial testing and follow-up was 26 months (range: 18-36 months).

#### Assessments

To assess ACE-R performance across all groups, we conducted a Bayesian ANOVA to test for group differences in the total scores, as well as in each cognitive sub-domain, followed by *post-hoc* independent samples *t*-test.

A set of further neuropsychological assessments was administered to all participants including the Cambridge Semantic Battery (CSB),^31^ the comprehension of spoken sentences subtest of the Comprehensive Aphasia Test (CAT),^32^ the forward and backward digit span from the Wechsler Memory Scale – Revised,^33,34^ the Trail Making Test,^35^ the Raven’s Coloured Progressive Matrices (CPM) test of non-verbal reasoning,^36^ and the Brixton test.^37^

Specific verbal and non-verbal episodic memory tests included the RAVLT^38,39^ and ROCF,^40^ respectively. The RAVLT consists of the following phases in order: (i) five learning trials of List A (Trials 1 through 5), consisting of 15 words; (ii) single trial of a new List B, also known as the interference list recall trial or Trial 6, consisting of 15 words; (iii) immediate recall, where the participant is asked immediately to recall as many words as possible from the repeated List A; (iv) delayed recall, where the participant is asked to recall as many words as possible from the List A after a 30-minute delay; and (v) 30-word recognition trial, where the participant is asked to respond by saying “yes” or “no” to words they recognize from List A and 15 distractor words that were neither on List A nor List B. For the recognition trial, the examiner read aloud each word while visually presenting the recognition page and pointing to the corresponding printed word. The ROCF consists of the following test conditions: (i) copy, where the participant is asked to copy the presented figure; (ii) immediate recall, where the participant is asked to draw the figure from memory after a short 3-minute delay; (iii) delayed recall, where the participant is asked to draw the figure from memory after a 30-minute delay; and (iv) recognition trial, where the participant is asked to respond by saying “yes” or “no” to parts of the figure they recognize from an array of 15 correct and 15 distractor/incorrect images. For the ROCF recognition trial, the examiner visually presented the recognition page, pointing to one image at a time.

#### Neuroimaging

As detailed in Henderson *et al*.,^29^ all participants (12 healthy controls, 27 patients) completed a T1-weighted 3T structural MRI scan on a Siemens PRISMA at the University of Cambridge (GRAPPA acceleration factor = 2) within 6 months of cognitive testing. Thirty-two participants were scanned at the MRC Cognition and Brain Sciences Unit with the following parameters: sagittal image acquisition, no. slices = 208, TR = 2000ms, TE = 2.85ms, flip angle = 8°, FOV = 228.8 x 281.6 x 281.6mm^3^, resolution matrix = 208 x 256 x 256, voxel size = 1.1 x 1.1 x 1.1mm^3^. Four participants were scanned at the Wolfson Brain Imaging Centre with the following closely-matched parameters: sagittal image acquisition, no. slices = 208, TR = 2000ms, TE = 2.93ms, flip angle = 8°, FOV = 228.8 x 281.6 x 281.6mm^3^, resolution matrix = 228.8 x 256 x 256, voxel size = 1.1 x 1.1 x 1.1mm^3^. T1-weighted MPRAGE images were pre- processed using Computational Anatomy Toolbox 12 (CAT12) (https://www.neuro.uni-jena.de/cat/) in the Statistical Parametric Mapping software (SPM12: Wellcome Trust Centre for Neuroimaging, https://www.fil.ion.ucl.ac.uk/spm/software/spm12/). A standard pre- processing pipeline was implemented with: (i) denoising, resampling, bias-correction, affine registration, and brain segmentation into three tissue probability maps (grey matter, white matter, cerebrospinal fluid); (ii) normalisation and registration to the Montreal Neurological Institute (MNI) template, (iii) smoothing using 8mm full-width-half-maximum (FWHM) Gaussian kernel. Segmented, normalised, modulated, and smoothed grey matter images were used for our region of interest (ROI) analysis. Of the nine patients who were followed longitudinally, six completed T1-weighted 3T structural MRI scans using the same parameters, detailed above.

Our *a priori* ROIs included 14 regions within the language and memory network derived from Harvard-Oxford cortical maps via FSLView: left and right hippocampus, posterior superior temporal gyrus (pSTG), posterior middle temporal gyrus (pMTG), Heschl’s gyrus, supramarginal gyrus (SMG), angular gyrus (AG), as well as bilateral posterior cingulate cortex (PCC) and precuneus. Each ROI was thresholded at >20% and binarised. Grey matter volumes in our ROIs were quantified as the number of grey matter voxels in each ROI controlling for age and total intracranial volume as nuisance covariates.

#### Statistical analysis

To test for group differences in demographics, we conducted a Bayesian ANOVA followed by a *post hoc* test for pairwise multiple comparisons. To assess ACE-R performance across all groups (i.e., controls, tAD, lvPPA, lvPPA+), we conducted a Bayesian ANOVA to test for group differences in the total scores, as well as in each cognitive sub-domain, followed by post *hoc* analyses.

To derive a weighted average of many cognitive domains and neuropsychological measures, we used principal component analysis to best summarise the cognitive measures. To maintain adequate sampling adequacy, we selected the following exemplar tests from a large neuropsychological battery that captured the primary language difficulties in lvPPA, memory deficits in tAD, as well as tasks with greater executive demands: CSB naming subtest, CAT comprehension of spoken sentences subtest, digit span backward, RAVLT and ROCF (i.e., immediate recall, delayed recall), Trail Making Test B, Brixton, and Raven’s CPM. We first imputed the missing data (7% overall) using estim_ncpPCA, a widely used method to impute with cross-validated principal component analysis.^41,42^ Scores on these eight tests were entered into a varimax-rotated principal component analysis. After selecting two components using parallel analysis, principal component scores were extracted for each individual participant. The principal component analysis was computed on only the first round, namely the cross-sectional patients’ data. Principal component scores were derived from the same principal component analysis model for controls and patients with a follow up visit to allow for between and within group comparisons. For each principal component, we tested the differences between group (i.e., controls, tAD, lvPPA, lvPPA+) using a one-way Bayesian ANOVA.

To assess verbal learning and recall using RAVLT scores, we conducted a 4 phase (early learning, late learning, immediate recall, delayed recall) x 4 group (control, tAD, lvPPA, lvPPA+) Bayesian ANOVA. Consistent with prior studies,^43^ Trials 1 and 2 were averaged to represent “early learning” and Trials 4 and 5 were averaged to represent “late learning”. Recognition was further assessed with a 2 condition (true positive, false positive) x 4 group Bayesian ANOVA. To assess visual learning and recall using ROCF scores, we conducted a 3 phase (copy, immediate recall, delayed recall) x 4 group (control, tAD, lvPPA, lvPPA+) Bayesian ANOVA. Recognition was assessed with 2 condition (true positive, false positive) x 4 group Bayesian ANOVA. We conducted *post hoc* Bayesian *t*-tests to test for group differences within each phase and condition.

For our imaging analyses, Bayesian linear regression assessed the relationship between principal component scores and grey matter volumes in the ROIs in patients, and separately across all groups including controls.

Given the relatively small number of patients with a follow-up visit, we analysed the longitudinal behavioural data in two ways. First, using the results of the principal component analysis, we visually presented the individual-level longitudinal variations capturing patient-specific movement along and across the principal component axes. Second, we employed a Bayesian approach to test whether each patient’s principal component scores and cognitive performance were significantly different from their representative diagnostic group as the normative sample across the two time-points. These group sample data were derived from the initial assessment scores with the sample size being 10 for lvPPA, eight for lvPPA+, and nine for tAD. Using the point and interval estimates of effect sizes for single case to normative sample design in neuropsychology,^44,45^ we derived a point estimate of the effect size for the difference between each patient and the group sample with an accompanying 95% credible interval, as well as a point and interval estimate of the abnormality of the patient’s score. This method made it possible to compare a particular participant’s principal component and cognitive scores to those of their diagnostic group average at initial assessment and at follow-up visit.

All Bayesian analyses were conducted in JASP version 0.18.3.0 with secondary frequentist statistics using RStudio v.2.4.0. Bayesian evidence in favour of the alternate hypothesis was interpreted using standard thresholds with the Bayes factor (BF): moderate (3 < BF < 10), strong (10 < BF < 30), very strong (30 < BF < 100), and extreme (100 < BF). BF of less than 0.33 provided evidence for the null hypothesis. Of note, 0.33 < BF < 3 implies the lack of sufficient precision (cf. power) to draw inferences either in favour of the null or alternative hypothesis.

## Results

### Part 1. Retrospective study of lvPPA, PCA and tAD

#### Demographics

As expected, there was evidence for a difference in age in patients (*P* < .001, BF > 100) which was driven by patients with PCA (mean = 58.51, SD = 12.05) being younger than those with tAD (mean = 68.25, SD = 8.99) and lvPPA (mean = 71.07, SD = 5.95). There was no evidence of a difference in age between tAD and lvPPA patients (*ns,* BF = 1.14). Bayesian ANOVA revealed no evidence of a difference in gender between groups (*ns*, BF = 0.58).

#### ACE-R

Figure 1A shows comparison data from research participants (*n* = 413) with a diagnosis of typical, amnesia-led AD (tAD) or AD-subtype (lvPPA and PCA) previously seen at memory clinics of the Cambridge University Hospitals. With a simplistic view from a categorical model, each subgroup is expected to be impaired in their core symptom but not in other cognitive domains. Thus, separate clusters would be expected. As shown in Figure 1A, neither of these two patterns was found. Instead, each group tended to be worse at their primary or core domain of impairment (see group ANOVAs below) but many cases had impairments in other “non-core impairment” domain areas. For example, many lvPPA and tAD patients showed impaired performance on both language and memory domains, as shown in the left scatterplot of Figure 1B. While most PCA patients were relatively preserved in language (as shown in the yellow dots populated in the right edge of the right scatterplot of Figure 1A), some patients were also impaired on language and memory.

**Figure 1.**
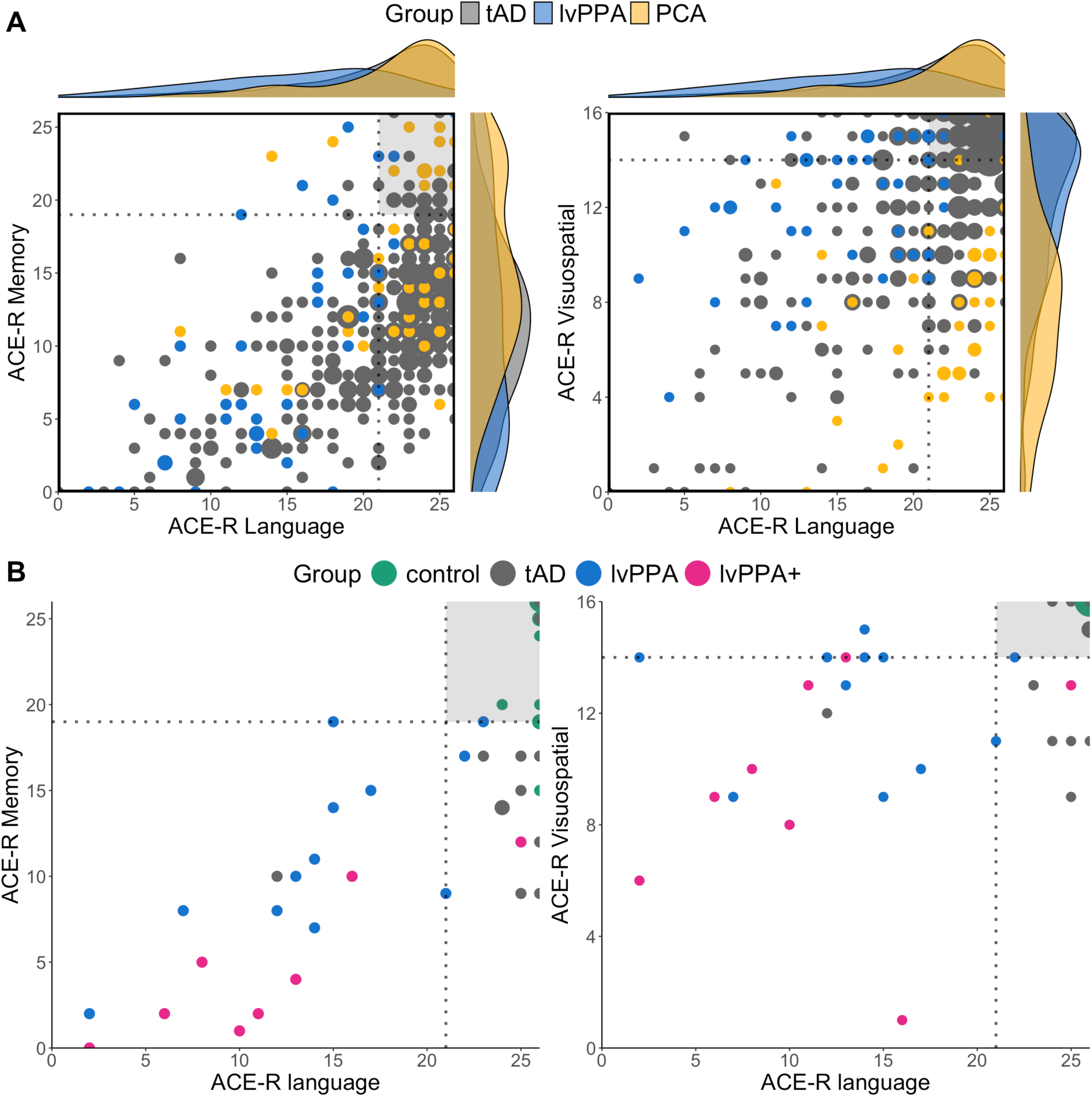
Scatterplots of ACE-R language, memory, and visuospatial domain scores over the (A) retrospective study population (tAD *n* = 329, lvPPA *n* = 44, PCA *n* = 40) and (B) prospective study population (control *n* = 12, tAD *n* = 9, lvPPA *n* = 10, lvPPA+ *n* = 8). The dotted lines represent the ACE-R cutoff scores from published healthy control normative data (Mioshi et al., 2006) and the shaded grey sections portray the quadrant of normality. In the top scatterplots, the larger dots portray more participants who had the same scores and the accompanying raincloud plots on the sides show the density curves of the data distribution per group.

Given the overlapping and non-discrete plots described above, one might wonder if this is simply a result of co-occurring deficits with increasing severity. Thus, it is informative to look at the relative proportion of patients with deficits in “core” *versus* “non-core” domains, stratified by total ACE-R scores as a measure of global severity. Table 1 shows the number of participants in each diagnostic group stratified by healthy control normative cut-off ranges. Despite the differences in sample size, all groups showed relatively equivalent distribution across the control range and cut-off ranges with around 9% of tAD, 15% of PCA, and 9% of lvPPA patients performing within the normal range (i.e., Groups 1 and 2) in terms of total ACE-R and the majority of patients falling below the cut-off ranges.

**Table 1.** Retrospective research participants stratified by ACE-R total scores.

| Grouped by ACE-R total scores | tAD (*n* = 329) | PCA (*n* = 40) | lvPPA (*n* = 44) |
| --- | --- | --- | --- |
| Group 1: Upper limit of normal range, i.e., within 1 SD of the control mean [scores of 89 and up] | 9 (2.74%) | 2 (5%) | 2 (4.55%) |
| Group 2: Lower limit of normal range, i.e., between 1 and 2 SD of the control mean [$85 \leq \text{score} < 89$] | 19 (5.78%) | 4 (10%) | 2 (4.55%) |
| Group 3: Below cut-off, i.e., 3 SDs below the control mean [$77 \leq \text{score} < 85$] | 58 (17.63%) | 5 (12.5%) | 2 (4.55%) |
| Group 4: Significantly below cut-off, i.e., more than 3 SDs below the control mean [scores of less than 77] | 243 (73.86%) | 29 (72.5%) | 38 (86.36%) |

Figure 2 shows the proportion of patients who were within the normal range and below the cut- off in each of ACE-R subdomains of cognition and highlights two notable observations: (1) even some mild patients who were within the normal healthy control range showed multi-domain cognitive impairments (e.g., deficits in attention and memory in addition to visuospatial in PCA Group 2 and deficits in visuospatial, attention and fluency in addition to memory in AD Group 2); and (2) the majority of patients belonged to Group 4 and showed prominent deficits in all cognitive domains.

**Figure 2.**
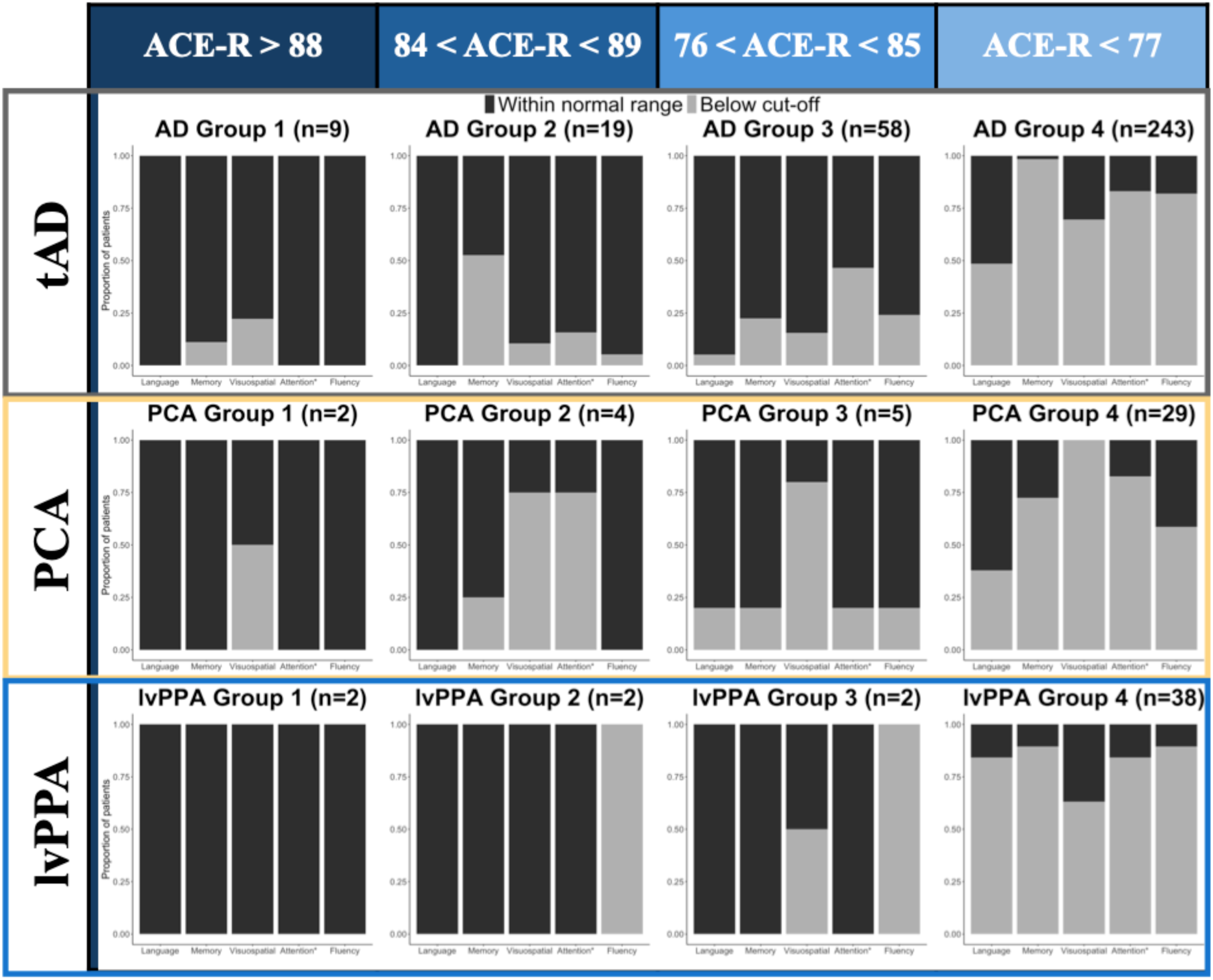
P**r**oportions **of patients who were within normal range or below published healthy control cut-off scores for each ACE-R cognitive domain stratified by overall performance.** The number of patients in each group is indicated in the title for each bar plot in curved brackets. Attention* denotes the ACE-R subdomain of attention/orientation. The four groups were stratified based on published cut-off values for the ACE-R: (1) Group 1 – upper limit of normal range (i.e., within 1 SD of the control mean); (2) Group 2 – lower limit of normal range (i.e., between 1 and 2 SD of the control mean); (3) Group 3 – below the cut-off (i.e., 3 SDs below the control mean); and (4) Group 4 – significantly below the cut-off (i.e., more than 3 SDs below the control mean).

Results of a Bayesian ANOVA revealed extreme evidence for a group effect on ACE-R language (F(2,410) = 18.00, *P* < 0.001, BF > 100) and visuospatial (F(2,410) = 24.31, *P* < 0.001, BF > 100) sub-scores. *Post hoc* analyses revealed extreme evidence for a difference between PCA relative to lvPPA and tAD patients (BF >100) for the visuospatial domain and between lvPPA relative to PCA and tAD patients (BF >100) for the language domain. Bayesian ANOVA revealed very strong evidence for a group effect on ACE-R memory (F(2,410) = 8.55, *P* < 0.001, BF = 85.15) driven by tAD and lvPPA patients having lower scores than PCA patients with evidence being extreme for the former (BF > 100) and moderate for the latter (BF = 4.62). *Post hoc* tests showed evidence for no difference between tAD (mean memory score = 11.28) and lvPPA (mean memory score = 11.18) patients (BF = 0.17). Results of a Bayesian ANOVA revealed anecdotal evidence (BF = 1.87) for the ACE-R total scores and no evidence (BF = 0.35) for the attention/orientation sub-scores.

### Part 2. Prospective study of lvPPA and tAD

#### Demographics

Table 2 shows the demographic details for the deep phenotype study of lvPPA compared with tAD and healthy controls. Bayesian ANOVA revealed evidence for no difference in all groups for age and handedness (*ns*, BF < 0.33) and there was no evidence for a difference in gender (*ns*, BF = 0.54) There was anecdotal evidence for a difference in symptom duration in patients (*ns*, BF = 1.66) which was driven by patients with tAD reporting longer symptom duration than those with lvPPA. There was anecdotal evidence for a difference in education (*ns*, BF = 1.91) which was driven by controls having higher levels of education than patients. However, *post hoc* tests did not reveal evidence for a difference between controls and each of the patient groups (*ns*, BF < 2.00), as well as between patients (*ns*, BF < 1.00).

**Table 2.**
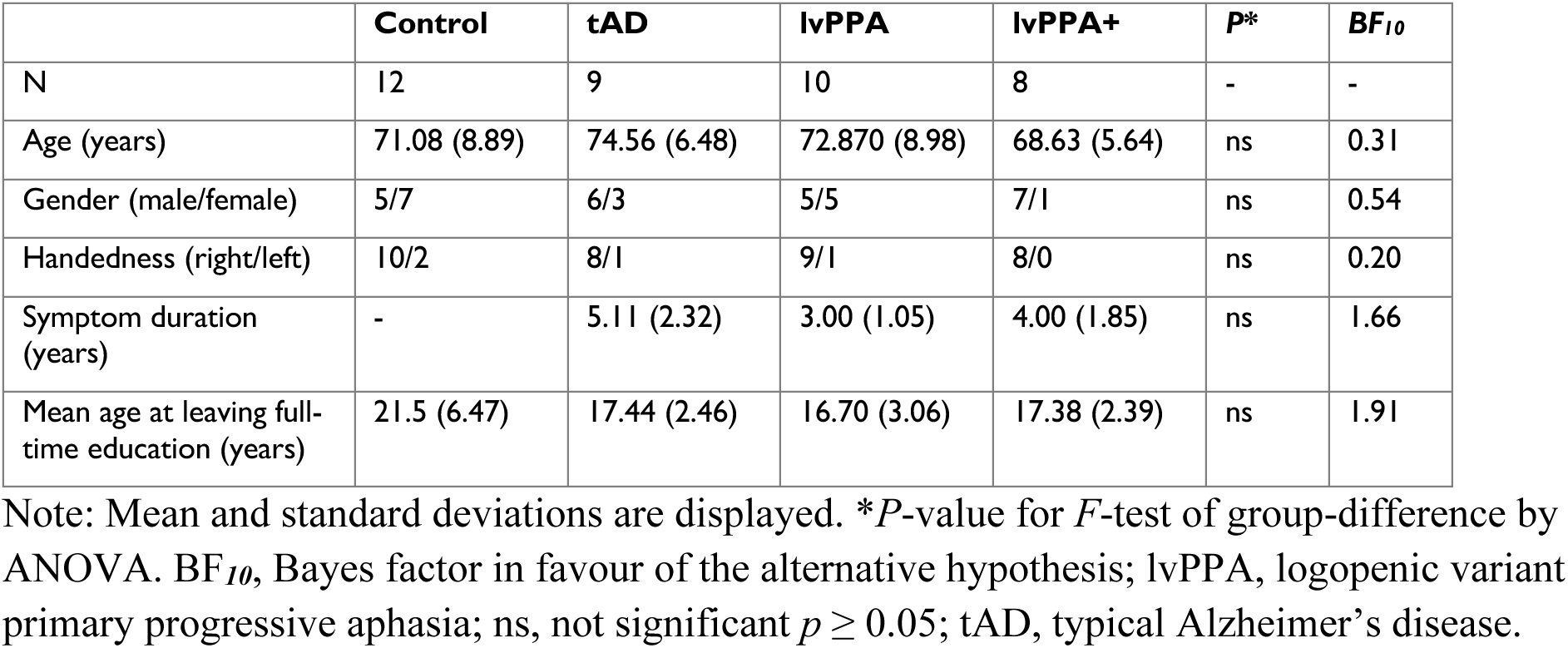
Demographics of the study cohort.

| | Control | tAD | lvPPA | lvPPA+ | $P^*$ | $BF_{10}$ |
| --- | --- | --- | --- | --- | --- | --- |
| N | 12 | 9 | 10 | 8 | - | - |
| Age (years) | 71.08 (8.89) | 74.56 (6.48) | 72.870 (8.98) | 68.63 (5.64) | ns | 0.31 |
| Gender (male/female) | 5/7 | 6/3 | 5/5 | 7/1 | ns | 0.54 |
| Handedness (right/left) | 10/2 | 8/1 | 9/1 | 8/0 | ns | 0.20 |
| Symptom duration (years) | - | 5.11 (2.32) | 3.00 (1.05) | 4.00 (1.85) | ns | 1.66 |
| Mean age at leaving full-time education (years) | 21.5 (6.47) | 17.44 (2.46) | 16.70 (3.06) | 17.38 (2.39) | ns | 1.91 |

Note: Mean and standard deviations are displayed. $P^*$ -value for $F$ -test of group-difference by ANOVA. $BF_{10}$ , Bayes factor in favour of the alternative hypothesis; lvPPA, logopenic variant primary progressive aphasia; ns, not significant $p \geq 0.05$ ; tAD, typical Alzheimer's disease.

#### Clinic-level assessment: ACE-R

Figure 1B visually summarises the relationship of ACE-R language versus memory and visuospatial sub-scores across all groups. Similar to the results of the retrospective analysis, each patient group tended to be worse at their primary or core domain of impairment (see group ANOVAs below) but (i) many cases had impairments in other “non-core impairment” domain areas and (ii) tAD and lvPPA groups performed similarly in the domain of memory.

Results of a Bayesian ANOVA revealed extreme evidence for a group effect on ACE-R total scores (F(3,34) = 48.91, *P* < 0.001, BF > 100). *Post hoc* analyses revealed extreme evidence for a difference between controls and all three patient groups (*P* ≤ 0.003, BF > 100). Across the patient groups, evidence for a difference in ACE-R total scores was extreme between tAD and lvPPA+ (*P* < 0.001, BF > 100), and strong between tAD and lvPPA (*P* = 0.007, BF = 24.72) and lvPPA and lvPPA+ (*P* < 0.001, BF = 12.69).

Results of Bayesian ANOVA revealed extreme evidence for a group effect on all ACE-R subdomain scores (*P* < 0.001, BF > 100). For ACE-R language sub-scores, *post hoc* pairwise multiple comparisons revealed extreme evidence for a difference between controls and patients with tAD versus those with lvPPA and lvPPA+ (*P* < 0.001, BF > 100). There was anecdotal evidence for a difference between controls and tAD patients (*P* = 0.95, BF = 2.02) and no evidence between lvPPA and lvPPA+ patients (*P* = 0.12, BF = 0.94).

For ACE-R memory sub-scores, evidence for a difference was extreme between controls and patients with lvPPA and lvPPA+ (*P* < 0.001, BF > 100), very strong between tAD and lvPPA+ (*P* < 0.001, BF = 80.15), and strong between controls and tAD (*P* = 0.004, BF = 29.01) and between lvPPA and lvPPA+ (*P* = 0.004, BF = 13.23). There was no evidence for a difference between tAD and lvPPA patients (*P* = 0.49, BF = 0.75).

For ACE-R visuospatial sub-scores, we found differences in controls versus patients only with evidence being extreme in controls relative to patients with lvPPA and lvPPA+ (*P* ≤ 0.05, BF > 100) and strong relative to patients with tAD (*P* = 0.11, BF = 15.01). The difference between controls and tAD patients was not corroborated by frequentist Tukey’s HSD for multiple comparisons.

As shown in Supplementary Fig. 1, evidence for a difference in ACE-R fluency sub-scores was extreme between controls and all patient groups (*P* < 0.001, BF > 100), strong between patients with tAD versus lvPPA+ (*P* = 0.001, BF = 17.22), and moderate between patients with tAD versus lvPPA (*P* = 0.02, BF = 5.95). For ACE-R attention and orientation sub-scores, we found differences in patients with lvPPA+ versus the rest of the groups with evidence being extreme relative to controls (*P* < 0.001, BF > 100) and very strong relative to patients with tAD and lvPPA (*P* < 0.001, BF > 30). Evidence for a difference between controls and lvPPA patients was strong (BF = 42.43) but this comparison was not corroborated by frequentist *post hoc* Tukey’s HSD multiple comparisons (*P* = 0.09).

#### Detailed neuropsychology

##### Neuropsychological principal component analysis

Scores on eight neuropsychological tests per participant were entered into a principal component analysis with varimax rotation. Two principal components explained 60.4% of the variance (Kaiser-Meyer-Olkin = 0.65; Bartlett’s Test of Sphericity *X*^2^ = 172.10, *df =* 45, *P* < 0.001). As shown in Supplementary Table 1, all tests except ROCF and digit span backward loaded heavily on principal component (PC) 1 and thus we labelled this PC as ‘multi-domain cognition’. Only ROCF (i.e., immediate recall, delayed recall) loaded heavily on PC 2 which we labelled as ‘non-verbal memory’. As shown in the group performance patterns in Figure 3, the results of Bayesian ANOVAs showed no evidence for a difference between tAD and lvPPA patients for both PC 1 and PC 2 (*ns*, BF < 2).

**Figure 3.**
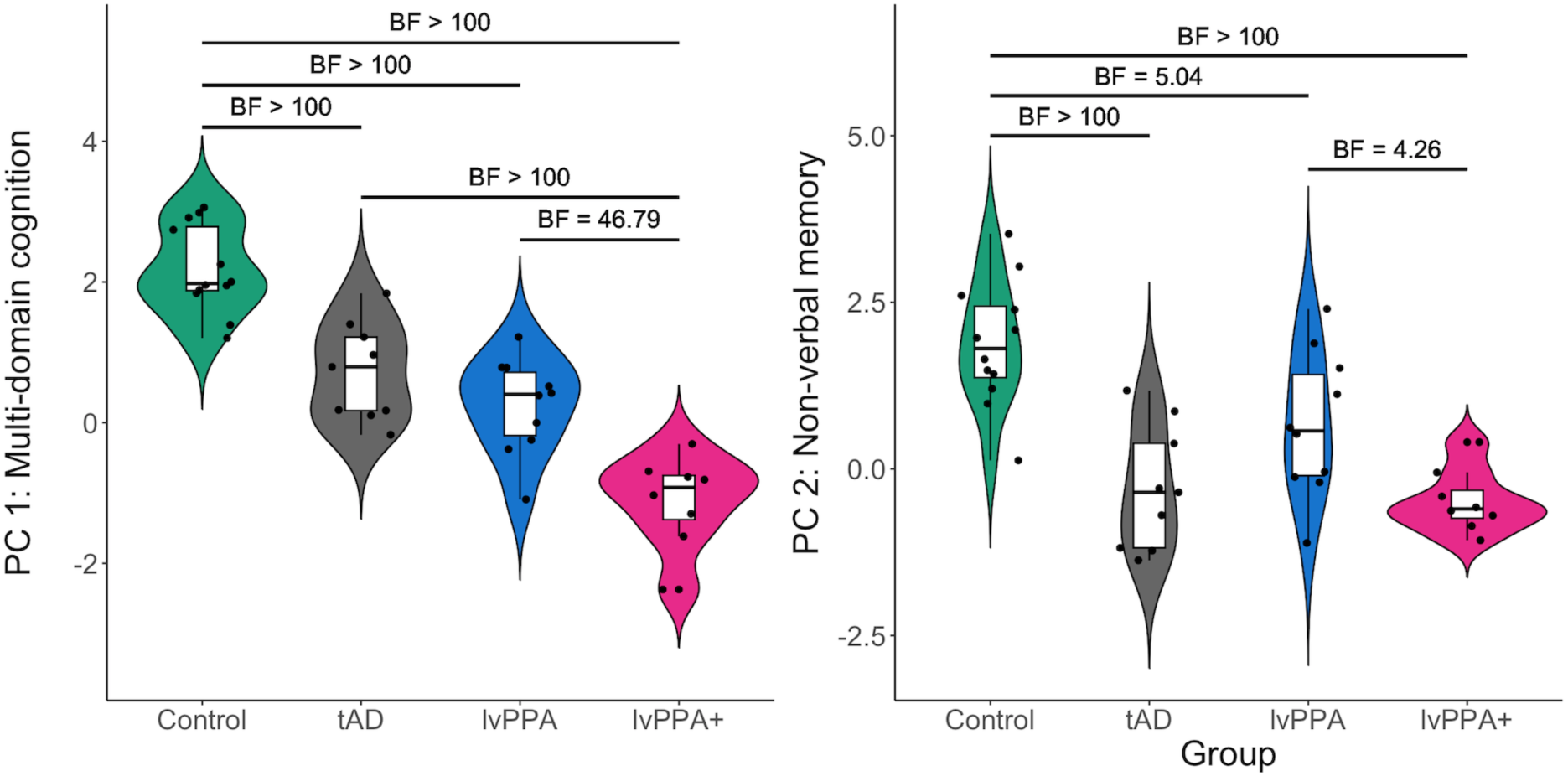
P**r**incipal **component analysis scores across diagnostic groups**. (**A**) Principal component (PC) 1: ‘multi-domain cognition’; and (**B**) PC 2: ‘non-verbal memory’. Bayes factors resulting from *post hoc* analyses are shown for each group comparison.

For PC 1, the results from a one-way Bayesian ANOVA revealed group differences (F(3,35) = 42.43, *P* < 0.001, BF > 100), driven by controls having higher PC 1 scores than all patients (*P* < 0.001, BF > 100). Additionally, evidence for a difference was extreme in tAD versus lvPPA+ patients (*P* < 0.001, BF > 100) and very strong in lvPPA versus lvPPA+ patients (*P* < 0.001, BF = 46.79).

For PC 2, a one-way Bayesian ANOVA revealed group differences (F(3,35) = 14.78, *P* < 0.001, BF > 100), driven by controls having higher PC 2 scores than all patients. Evidence was extreme in controls versus tAD and lvPPA+ patients (*P* < 0.001, BF > 100), and moderate versus lvPPA patients (*P* = 0.02, BF = 5.04). There was also moderate evidence for a difference in PC 2 scores between lvPPA and lvPPA+ patients (*P* = 0.05, BF = 4.26).

##### Episodic memory specific scores: RAVLT and ROCF

The detailed Bayesian analyses of the verbal and non-verbal memory testing are reported next but the key results for this analysis were that lvPPA patients (i) exhibited impaired performance on both verbal and non-verbal episodic memory tests compared to controls and (ii) performed similarly when compared to tAD patients (Figure 4).

**Figure 4.**
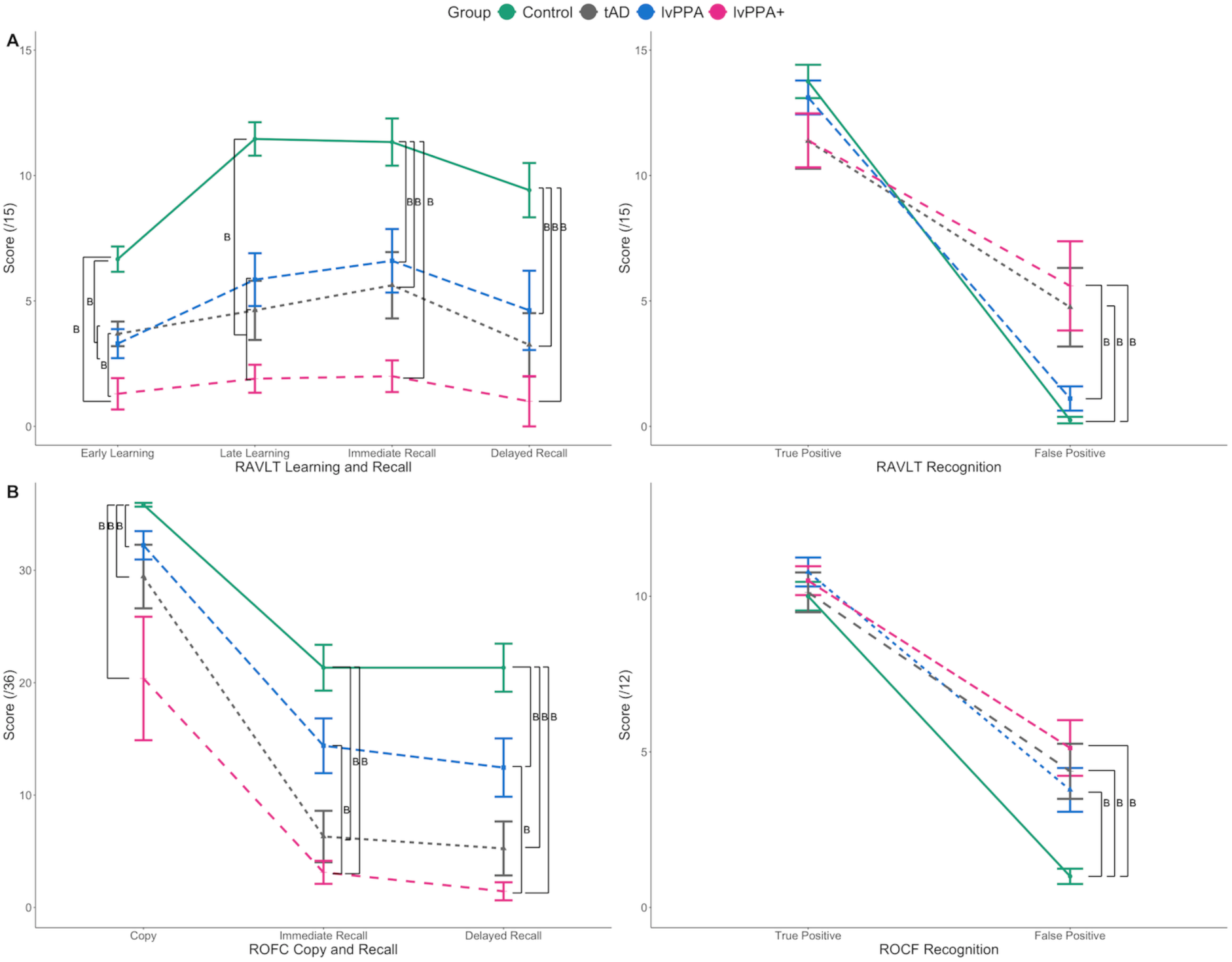
**(A) Rey Auditory Verbal Learning Test (RAVLT) and (B) Rey Osterrieth complex figure (ROCF) scores across groups**. The letter ‘B’ indicates bayes factors greater than 3 from all *post hoc* group comparisons. Of note, Trials 1 and 2 of the RAVLT were averaged to represent “early learning” and Trials 4 and 5 were averaged to represent “late learning”.

More specifically, to assess verbal memory performance, a 4 phase (early learning, late learning, immediate recall, delayed recall) x 4 group Bayesian ANOVA was conducted with RAVLT scores which revealed extreme evidence in favour of an effect of group (F(3,122) = 43.81, *P* < 0.001, BF > 100) and phase (F(3,122) = 5.24, *P* = 0.002, BF > 100). RAVLT recognition scores were assessed with 2 condition (true positive, false positive) x 4 group Bayesian ANOVA which revealed extreme evidence in favour of an effect of condition (F(1,60) = 206.61, *P* < 0.001, BF > 100) and group-by-condition interaction (F(3,60) = 8.91, *P* < 0.001, BF > 100), as well as very strong evidence for an effect of group (F(3,60) = 1.18, *P* = 0.32, BF = 66.28), for which the significance was not corroborated by frequentist ANOVA.

To assess non-verbal memory performance, a 3 phase (copy, immediate recall, delayed recall) x 4 group Bayesian ANOVA was conducted with ROCF scores, and we found extreme evidence in favour of an effect of group (F(3,100) = 33.58, *P* < 0.001, BF > 100) and phase (F(2,100) = 83.89, *P* < 0.001, BF > 100). ROCF recognition scores were similarly assessed with 2 condition (true positive, false positive) x 4 group Bayesian ANOVA which revealed extreme evidence in favour of an effect of condition (F(1,66) = 259.24, *P* < 0.001, BF > 100) and group (F(3,66) = 6.71, *P* < 0.001, BF > 100), as well as strong evidence for a group-by-condition interaction (F(3,66) = 4.26, *P* = 0.008, BF = 26.38).

Figure 4 visually summarises the results of *post hoc* group comparisons using Bayesian independent samples *t*-tests for each phase of learning and recall, as well as recognition condition for RAVLT (see 4A) and ROCF (see 4B). Details of *post hoc* group comparisons are shown in Supplementary Table 2.

#### Neuroimaging: Associations between principal component scores and grey matter volume

When analysing patients’ data only, we found either anecdotal or no evidence for a relationship between PC scores and all ROIs. We found moderate evidence for a relationship between PC 2 scores and right hippocampal grey matter volume (*β* = 0.45, *t* = 2.49, *P* = 0.02, BF = 3.13; Figure 5). We found either anecdotal or no evidence for a relationship between PC 2 scores and all other ROIs (0.33 < BF < 3).

**Figure 5.**
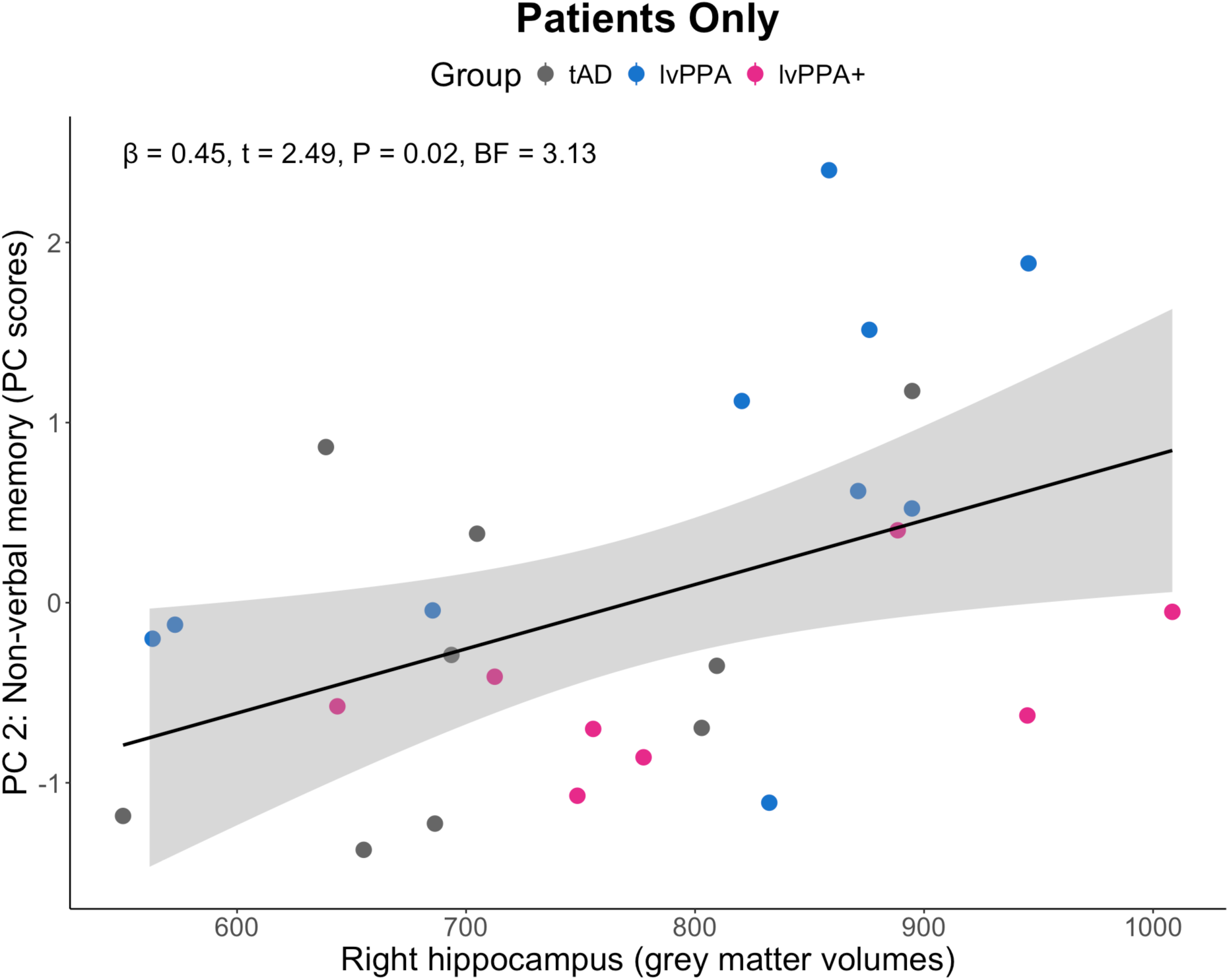
A**s**sociations **between Principal Component 2 scores and grey matter volume in the right hippocampus.** There was moderate evidence for a relationship between PC 2: “Non-verbal memory” scores and right hippocampal grey matter volumes (*β* = 0.45, *t* = 2.49, *P* = 0.02, BF = 3.13). BF, Bayes factor; lvPPA, logopenic variant primary progressive aphasia; tAD, typical Alzheimer’s disease.

In the whole group including controls, the results of Bayesian linear regression analyses revealed extreme evidence for the associations between PC 1 scores and grey matter volume in the left pMTG, pSTG, SMG, AG, and Heschl’s gyrus (BF > 100). In addition, evidence was very strong for the right pMTG (BF = 70.85) and bilateral PCC (BF = 30.89), strong for right pSTG (BF = 18.95), and moderate for the right AG (BF = 6.91), bilateral precuneus (BF = 4.87), and left hippocampus (BF = 4.10). Evidence for the association between PC 2 scores and grey matter volumes was extreme in the bilateral PCC, right pMTG, right Heschl’s gyrus, right pSTG, bilateral precuneus, left hippocampus and right AG (BF > 100), very strong in the right hippocampus (BF = 50.12), strong in the left SMG (BF = 25.77), left AG (BF = 23.36), left pMTG (BF = 21.56), right SMG (BF = 19.61) and left Heschl’s gyrus (BF = 10.35), and moderate in the left pSTG (BF = 9.99). Supplementary Fig. 2 shows the associations between PC scores and grey matter volume in the ROIs that showed extreme Bayesian evidence (BF > 100).

#### Longitudinal assessment

Figure 6 represents the longitudinal movements in PC scores of seven patients. The three tAD patients showed phenotypic dispersion, one (tAD1) remaining largely unimpaired and within the same category (i.e., quadrant of normality), another (tAD2) declining primarily in their core deficit of memory, and the third (tAD3) demonstrating relative preservation of non-verbal memory related PC scores but declining in multi-domain cognitive PC scores. Bayesian comparisons between the PC and available cognitive scores of each longitudinal case and their representative diagnostic group sample are shown in Supplementary Table 3. tAD3’s decline in global cognition, particularly in language, was corroborated by the Bayesian single case to a group sample comparison. tAD3’s ACE-R language, CSB naming and CAT comprehension of spoken sentences scores significantly declined in the follow-up visit, with a Bayesian two-tailed probability of < 0.001 that a member of the tAD group would obtain a score lower than the patient.

**Figure 6.**
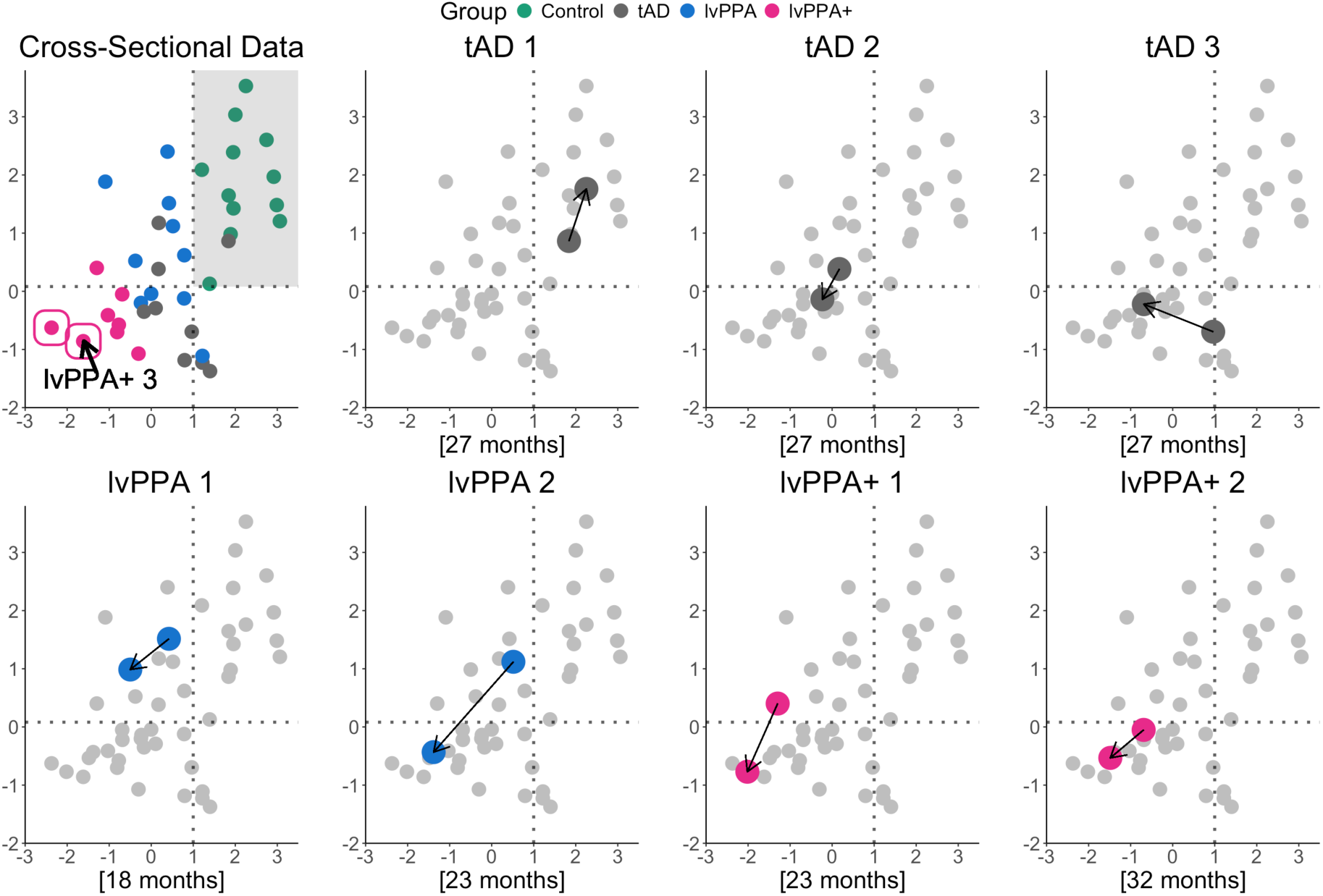
P**h**enotypic **variation in longitudinal patients within the principal component (PC) space.** The scatterplot on the top left visually summarises the principal component analysis results for all patients. The dotted lines indicate the 95% confidence interval of the mean of the healthy control PC scores and the grey shaded area shows the quadrant of normality. The two cases circled in pink could not complete any of the follow-up cognitive tasks. Of the two, case “lvPPA+ 3” completed a follow-up MRI scan as shown in Supplementary Fig. 3. Each longitudinal case is plotted individually with arrows indicating the change from initial to follow-up scores in each dimension. The square brackets delineate the time in months from initial to follow-up assessment.

The four lvPPA and lvPPA+ patients showed decline in both PC 1 and PC 2 scores. LvPPA2’s scores on language (ACE-R language), memory (ROCF recognition), visuospatial/executive functioning (Raven’s CPM), and global cognition (PC 1) at the follow-up visit significantly differentiated the case from the lvPPA group sample with a Bayesian two-tailed probability ≤ 0.05. Similarly, lvPPA1’s language (ACE-R language) and memory (ACE-R memory, RAVLT recognition) scores differentiated the case from the lvPPA group sample with a Bayesian two- tailed probability ≤ 0.05. In other words, as shown in the Bayesian point estimates, the percentage of the lvPPA group population falling below the two cases’ scores on these tests is found to be very low. Supplementary Fig. 3 shows the initial and follow-up MRI scans of six patients who were scanned at both time-points.

## Discussion

This study investigated whether AD phenotypes are distinct clinical entities or represent positions within a graded multidimensional space.^3,9,10,17^ First, we examined the comparative distributions of ACE-R performance in patients diagnosed with typical AD, lvPPA, and PCA using a large retrospective dataset of past research participants (*n* = 413). Next, in a prospective study of lvPPA compared to typical AD (*n* = 27), we addressed the following questions: (1) Is the multidimensional cognitive pattern of impairment only present in the advanced lvPPA cases and how do they compare to tAD? (2) Do memory deficits in lvPPA appear in a simple clinic-level cognitive assessment or require in depth research-level investigation? (3) To what extent is performance on verbal episodic memory attributable to language impairment? (4) Do the patterns of decline in lvPPA and typical AD stay categorical or multidimensional over time? In the imaging analyses, we explored the associations between principal component analysis derived scores and grey matter volumes in key memory- and language-related brain regions, at baseline and longitudinally. In the sections below, we revisit these questions, interpret our findings and note directions for future research.

### Is the multidimensional cognitive pattern of impairment only present in the advanced cases?

Our first question was whether the multidimensional cognitive pattern of impairment is only present in the advanced atypical AD cases and how they compare to that of tAD. Using the retrospective data of research participants, we examined the relative proportion of patients with deficits in “core” (i.e., memory in tAD, language in lvPPA, visual in PCA) *versus* “non-core” domains, stratified by total ACE-R scores as a measure of global severity. Two key findings emerged: (1) even some mild patients, who were within the normal healthy control range, showed multi-domain cognitive impairments (e.g., deficits in attention and memory in addition to visuospatial in PCA and deficits in visuospatial, attention and fluency in addition to memory in AD); and (2) the majority of patients showed prominent deficits in all ACE-R cognitive domains.

Prior studies have reported that visuospatial deficits may be present in tAD patients or emerge later as they progress to multi-domain dementia.^3,46^ The retrospective ACE-R data also showed that, regardless of diagnostic group, patients with advanced cognitive impairment (ACE-R significantly below the control cut-off) showed visuospatial deficits. The visuospatial weakness observed in all groups reflect the graded weightings of the features of the AD phenotype.^3^ It is worth noting that the ACE-R visuospatial results shown in Figure 1, where many tAD, most lvPPA, and all lvPPA+ patients had lower scores relative to controls, is in line with the difference between controls and all patient groups on the copy phase (i.e., visuoconstruction) of the non-verbal episodic memory task (see section below).

This multidimensional cognitive pattern of impairment was also present in the prospective study. The first component from the principal component analysis predominantly represented tests of language, visuospatial/executive functioning and verbal memory, and thus we labelled this PC as ‘multi-domain cognition’. While there were large expected differences on PC 1 between controls and all patient groups, and between tAD and lvPPA relative to lvPPA+ patients, we found no evidence that PC 1 multi-domain cognition scores differed between tAD and lvPPA patients. PC 2, predominantly represented non-verbal memory, also differentiated controls from all patients with, again, no differences between tAD and lvPPA, as well as tAD and lvPPA+ patients. These findings challenge the notion that lvPPA is a distinct nosologic entity.

### Do memory and other cognitive deficits appear in a simple clinic-level cognitive assessment in atypical AD phenotypes?

As a clinic-level assessment, the ACE-R is a useful contributor to the differential diagnosis of dementias with good specificity and sensitivity.^47–50^ We had a double opportunity to examine the comparative distributions of ACE-R performance across a full clinical spectrum of AD with the large dataset of past research participants (*n* = 413) and our prospective sample (*n* = 27).

The retrospective study revealed that multi-domain cognitive deficits across different AD phenotypes can be captured in a simple clinic-level assessment. There were three notable findings: (1) when stratified by overall ACE-R performance, even some mild patients who were within the normal healthy control range showed multi-domain cognitive impairments; (2) the majority of patients showed prominent deficits in multiple cognitive domains regardless of diagnostic group; and (3) whereas the prominent deficits in each diagnostic group were captured in the results of a Bayesian ANOVA (i.e., visuospatial in PCA, language in lvPPA, and memory in tAD), we found evidence for no difference between AD and lvPPA patients in the domain of memory.

The prospective analysis included patients who met stringent diagnostic criteria for both PPA and lvPPA,^12,27^ and thus we were able to compare these patients to typical amnestic- predominant AD, as well as the more progressed lvPPA+ patients. While the tAD group showed an amnestic predominant profile, the lvPPA and lvPPA+ groups exhibited impaired performance across language, memory and visuospatial domains when compared with controls. Consistent with the retrospective analysis, there was no evidence for a difference between tAD and lvPPA patients in the subdomain of memory. Taken together, these results highlight the graded distinctions (as shown as each group’s paradigmatic domain of impairment) and overlap that is evident between typical and atypical AD phenotypes.

### To what extent is performance on verbal episodic memory attributable to language impairment?

Because many screening tools and episodic memory tests are conducted using verbal materials, reduced memory scores in lvPPA could reflect the patients’ aphasia rather than amnesia per se.^51^ To investigate the extent to which memory test performance may be attributable to the patients’ aphasia, in our prospective study, we administered the most commonly used verbal and non-verbal episodic memory tests. Consistent with the ACE-R findings, lvPPA patients were impaired relative to controls in all aspects of non-verbal learning and recognition on the ROCF, as well as the RAVLT. Importantly and strikingly, lvPPA and tAD patients showed the same pattern of impairment across both tests. Moreover, this similar pattern of memory impairment was apparent in the clinic-level ACE-R (in both prospective and retrospective data) and in specific neuropsychological tests of episodic memory across verbal and non-verbal domains.

To summarise, there were two key findings: (1) the results of our detailed assessments, clinic-level and specific memory tests all consistently showed that multi-domain cognitive impairments are present in lvPPA patients across disease severity, as well as in tAD patients to a lesser extent when compared to controls; and (2) lvPPA patients’ impaired performance on episodic memory, as well as the lack of a difference when compared to tAD patients, suggest that memory deficits are present even in some mild lvPPA patients. This latter finding is consistent with prior studies that have shown that multi-domain cognitive deficits such as verbal memory, visuospatial functioning, spatial orientation and working memory, are present in lvPPA patients even when controlling for aphasia severity.^52–56^

### Do the patterns of decline in lvPPA and typical AD stay categorical or multidimensional over time?

Longitudinal investigations have immense value in addressing the gaps in our understanding about the trajectory of neurocognitive decline in AD. Given the challenges of this type of research, it is perhaps not surprising that such studies are rare; but they are needed to determine whether, with progression, AD phenotypes remain identifiably distinct or gradually merge. Our longitudinal tracking of a subset of patients provided insights about the phenotypic dispersion in tAD and lvPPA cases. In the three tAD patients, one remained largely preserved in higher cognition, another progressed more within the domain of non-verbal memory (in other words, staying in the same category), and the third exhibited significant global cognitive impairments, particularly in the language domain, over time. The four lvPPA and lvPPA+ patients showed declines in PC 1 ‘multi-domain cognition’ scores, which were loaded by tests of language, visuospatial/executive functioning, and verbal memory, as well as in PC 2 ‘non-verbal memory’ scores. As shown in Supplementary Table 3, declines in specific memory, language and/or visuospatial test scores were also evident. The significant overlap between different phenotypes in the bottom left quadrant of the top left scatterplot of Figure 6, as well as five out of seven patients occupying this space over time, strongly signals that there are graded variations within and between the AD phenotypes, further strengthening the multidimensional hypothesis.^3,9,11^

### Neuroimaging

Using a region of interest (ROI) approach, we explored the associations between PC scores and grey matter volume in memory- and language-related brain regions. In the whole group, we found extreme evidence for the associations between PC 1 ‘multi-domain cognition’ scores and grey matter volumes in the language network that are typically atrophied in lvPPA, namely the left pMTG, pSTG, SMG, AG, and Heschl’s gyrus, as well as strong evidence for regions reported to be crucial for episodic memory encoding and retrieval, such as the left hippocampus and bilateral precuneus, and verbal working memory including temporo-parietal regions.^10,57–61^

PC 2 ‘non-verbal memory’ scores were also found to be associated with grey matter volumes in the bilateral hippocampi, posterior cingulate, and temporo-parietal regions, commensurate with their roles in recall memory, spatial attention and episodic memory.^62–64^ When excluding controls, the only evidence for the association between principal component scores and grey matter volumes in key language and memory regions was found between PC 2 scores and right hippocampal grey matter volumes as shown in Figure 5. In our previous study,^29^ whereas the lvPPA group showed an asymmetrical pattern of reduced grey and white matter intensity in the language-dominant left hemisphere, including a significant portion of the lateral and medial temporal lobe, the more progressed lvPPA+ patients showed right mediotemporal grey and white matter reductions. The relationship between right hippocampal volumes and non-verbal memory-related PC scores is noteworthy in the context of the emergence of episodic memory deficits in lvPPA. Mesulam *et al.* ^5^ proposed that the preservation of episodic memory in lvPPA might be due to the unilaterality of mediotemporal degeneration. One proposal would be that episodic memory deficits, while present even in mild lvPPA patients, becomes more prominent in clinical presentation when atrophy spreads bilaterally over time to include right mediotemporal regions. Of course, such a proposal mirrors the classic neurosurgical data from patient HM and others.^65,66^

### Limitations

First, our sample size for AD confirmatory status with blood biomarkers, as well as the longitudinal patient data, is relatively small. However, clinico-pathological correlations are high in lvPPA. Second, our longitudinal study size was smaller than intended, in large part due to effects of the COVID-19 pandemic on participants and research infrastructure, but these patients are rare and longitudinal investigations are consequently uncommon in atypical AD samples. Third, for our retrospective dataset, clinical details, other assessments and neuroimaging data were not available. Despite the lack of details such as symptom duration and clinical dementia ratings, this large dataset provided a frame of reference to support the study’s broader aims. We further mitigated the lack of such measures by tabulating performance by total ACE-R scores as a proxy for cognitive severity. Importantly, direct comparisons of cognitive performance in typical and atypical AD phenotypes are rare, and thus our findings question the classical view of AD phenotypes encapsulated within distinct, categorical boundaries.

## Conclusion

The graded distinctions amongst typical amnestic and atypical (language and visual) phenotypes of AD support a transdiagnostic, multidimensional neurocognitive geometry proposal for those sharing the same underlying AD disease process. The findings from the current study are particularly relevant in the context of clinical management and treatment. Large-scale AD cohort studies tend to focus on typical, amnestic presentations of AD, excluding lvPPA and other AD phenotypes like PCA. Given the progress towards effective treatments that aim at the pathology, irrespective of clinical diagnoses, inclusion of all AD phenotypes is critical with a particular focus on transdiagnostic symptoms, considering their relevance to disease burden and interventions. This study provides supportive evidence for the possible future use of anti-amyloid monoclonal antibodies for atypical forms of AD. Improved clinical characterisation across disease severity may help people affected by AD and their clinicians to set expectations, manage symptoms, and initiate support.

## Data availability

The authors confirm that anonymised data supporting the findings of this study are available within the article and its Supplementary material.

## Acknowledgements

We sincerely thank our patients and their families for supporting this work.

## Supplementary material

**Supplementary Table 1.** Loadings for neuropsychological battery principal component analysis.

| **Measure** | **PC 1 (“Multi-domain cognition”)** | **PC 2 (“Non-verbal memory”)** |
| --- | --- | --- |
| CSB Naming | **0.70** | -0.10 |
| CAT Comprehension of Spoken Sentences | **0.89** | 0.00 |
| Trail Making Test B | **0.84** | 0.32 |
| Brixton | **0.52** | 0.20 |
| Raven’s Coloured Progressive Matrices | **0.61** | 0.26 |
| Digit Span Backward | 0.40 | 0.22 |
| RAVLT Immediate Recall | **0.74** | 0.17 |
| RAVLT Delayed Recall | **0.69** | 0.14 |
| ROCF Immediate Recall | 0.13 | **0.98** |
| ROCF Delayed Recall | 0.17 | **0.96** |

Rotation: Orthogonal varimax. Loadings above a threshold of 0.5 are bolded. PC, principal component; TTR, type-to-token ratio. CAT, Comprehensive Aphasia Test; CSB, Cambridge Semantic Battery; RAVLT, Rey Auditory Verbal Learning Test; ROCF, Rey-Osterrieth Figure Copy.

**Supplementary Table 2.**
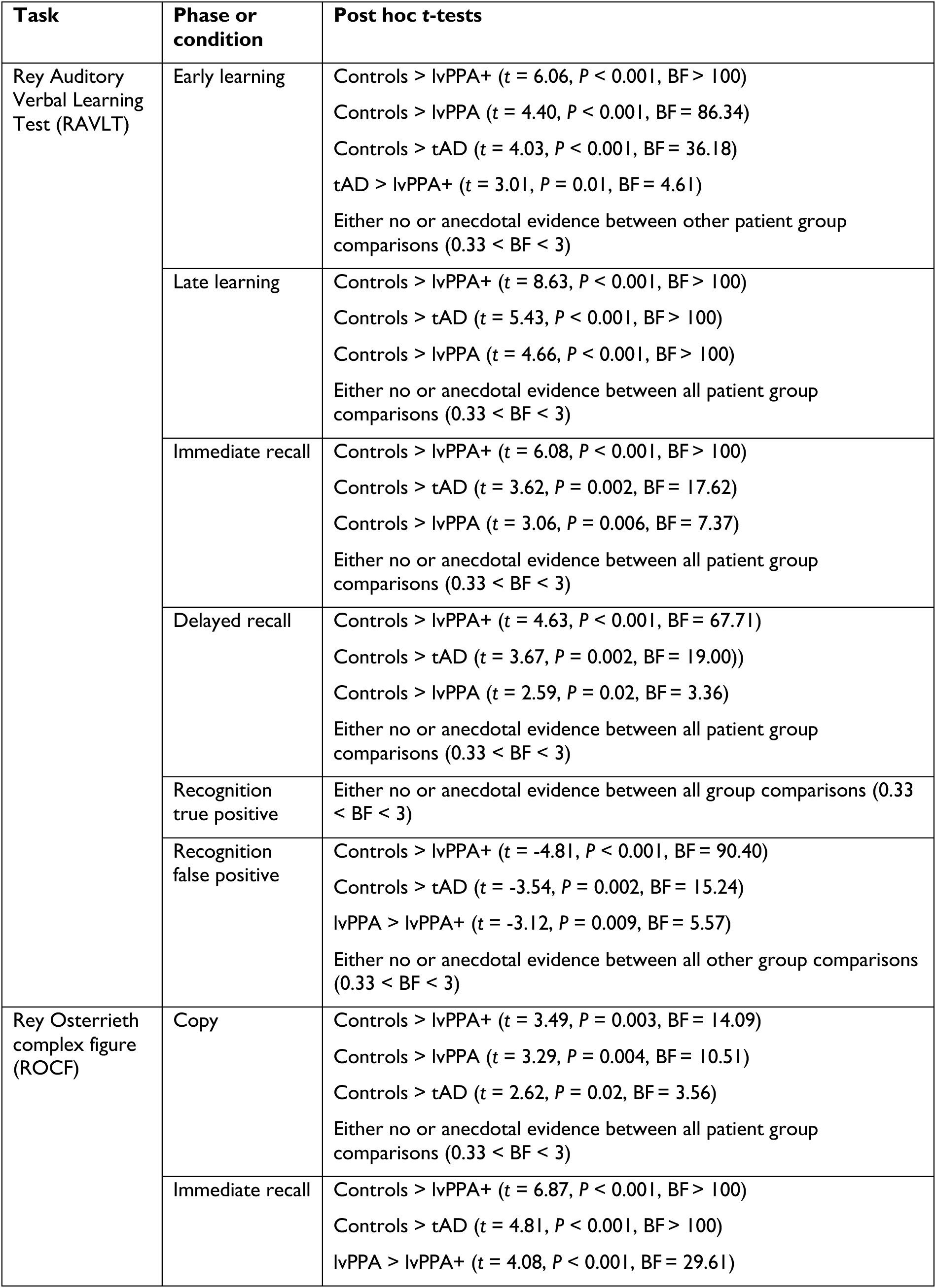
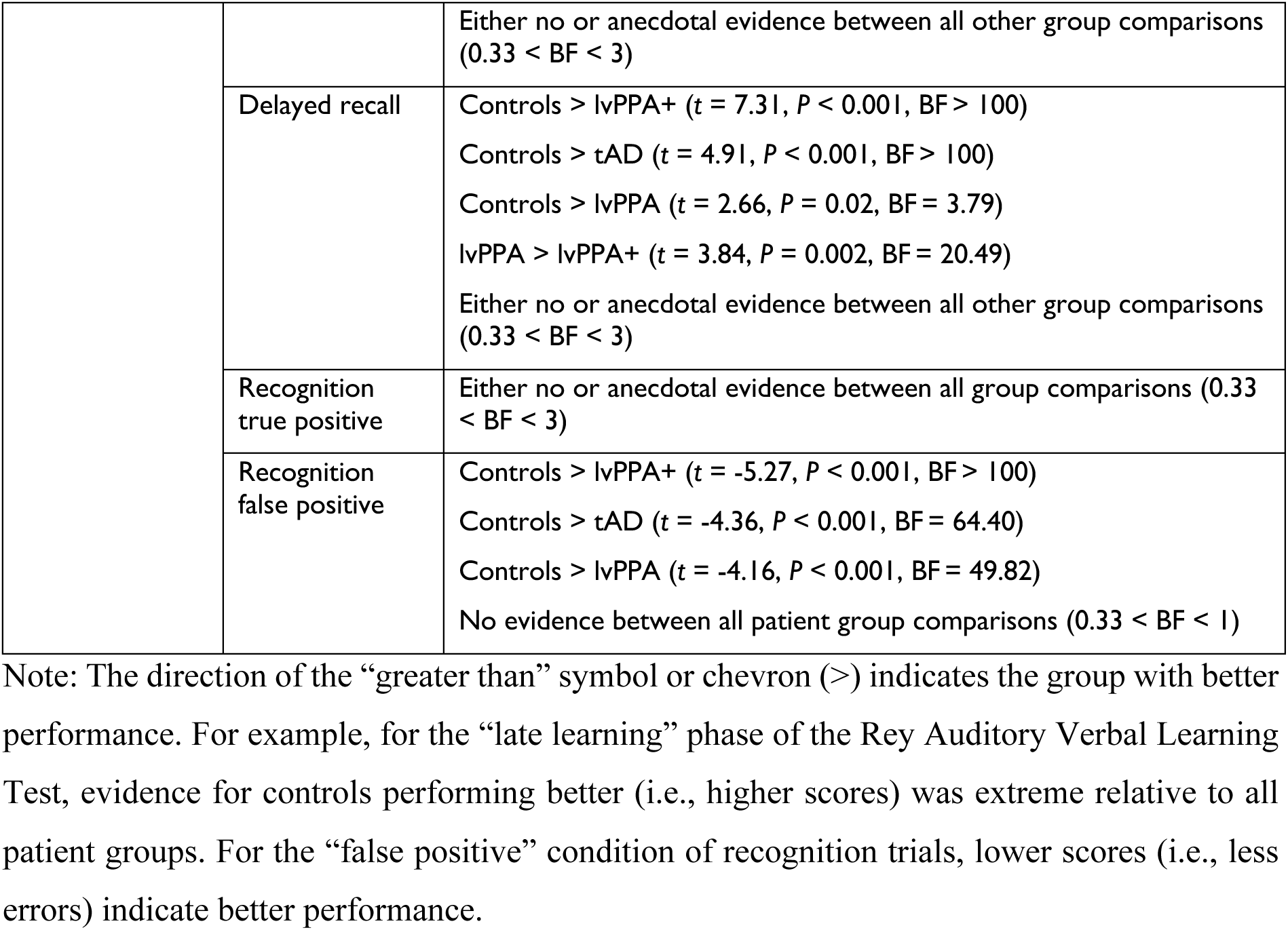
Verbal and non-verbal episodic memory tests *post hoc* group comparisons.

**Supplementary Table 3.**
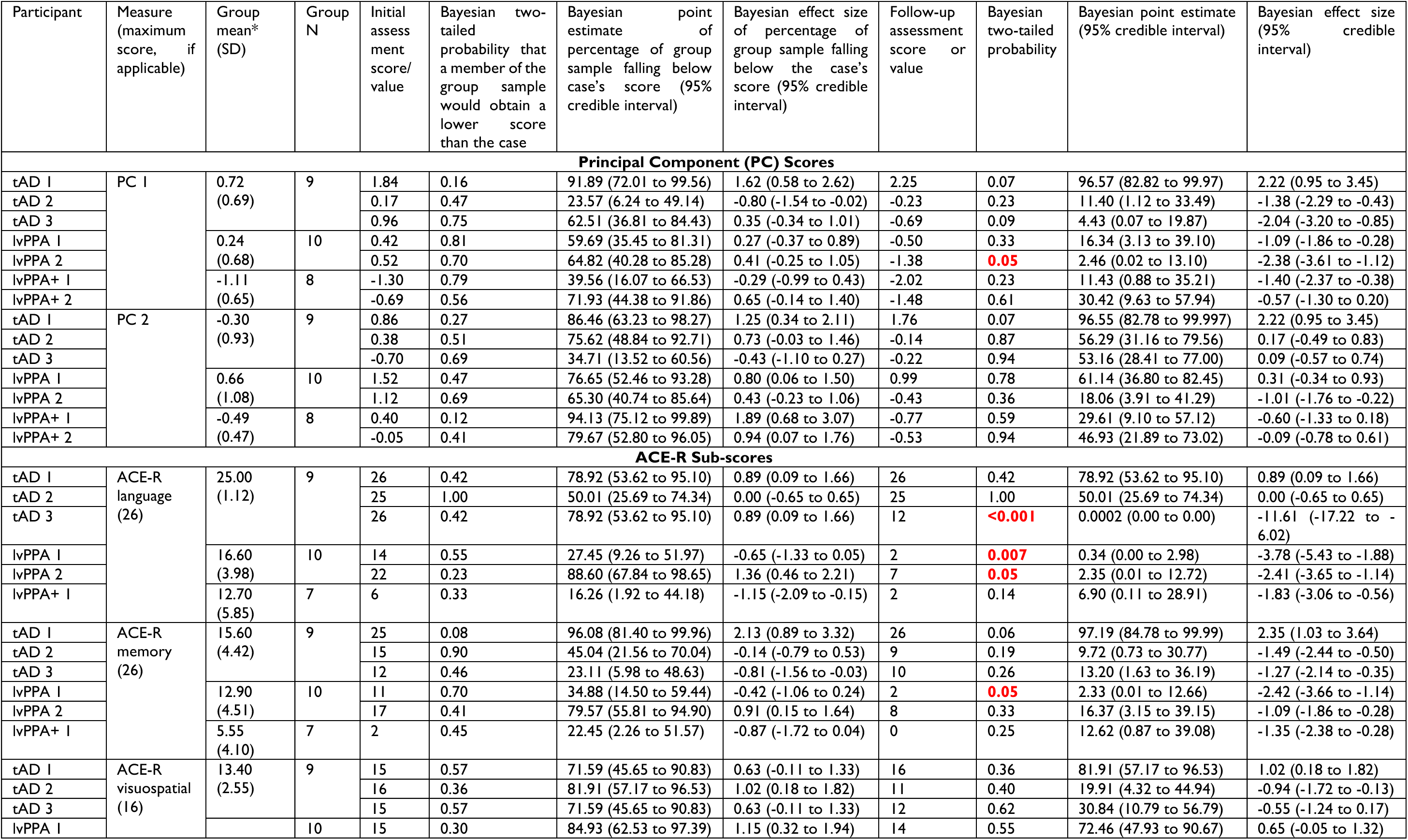
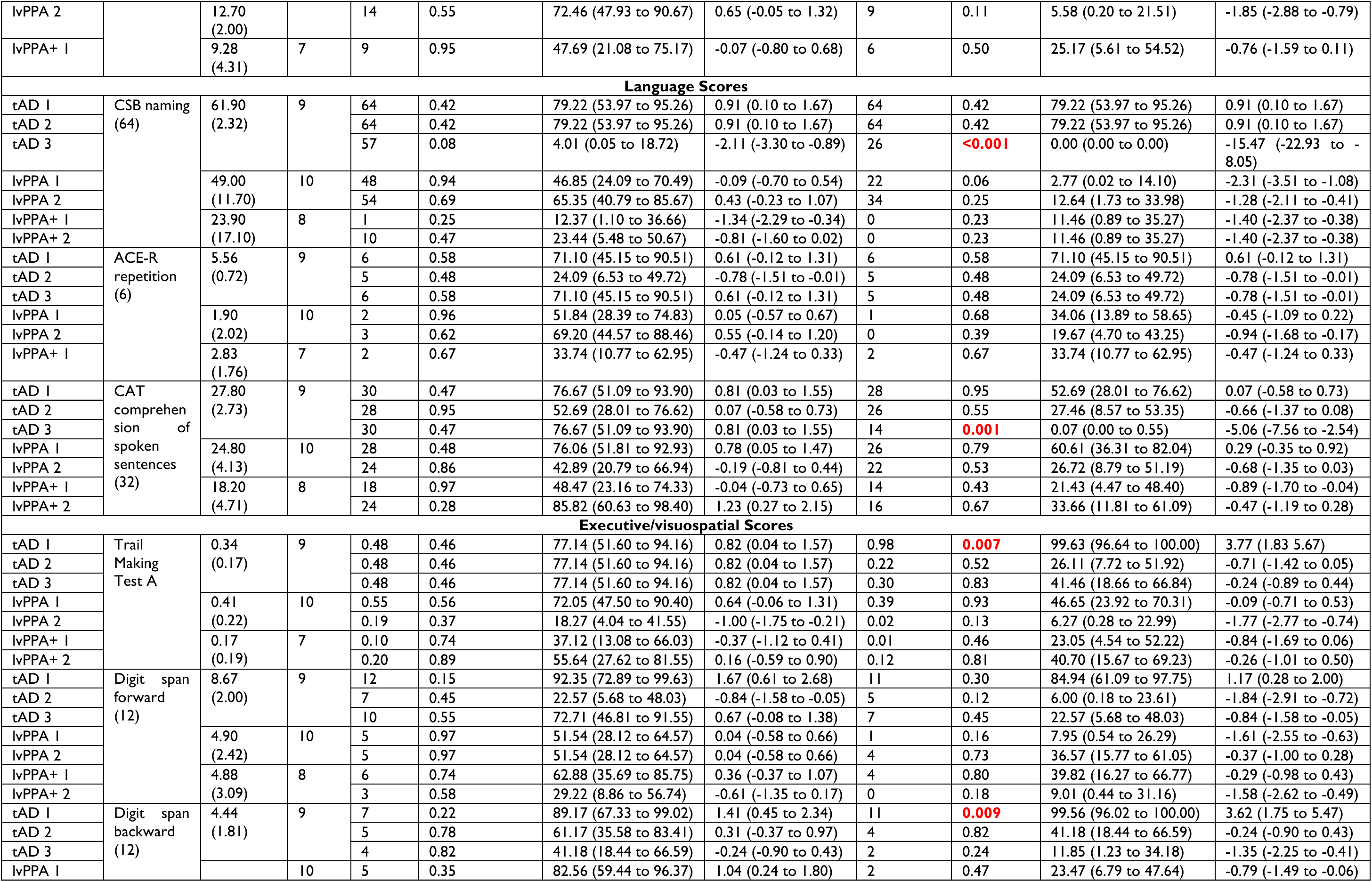
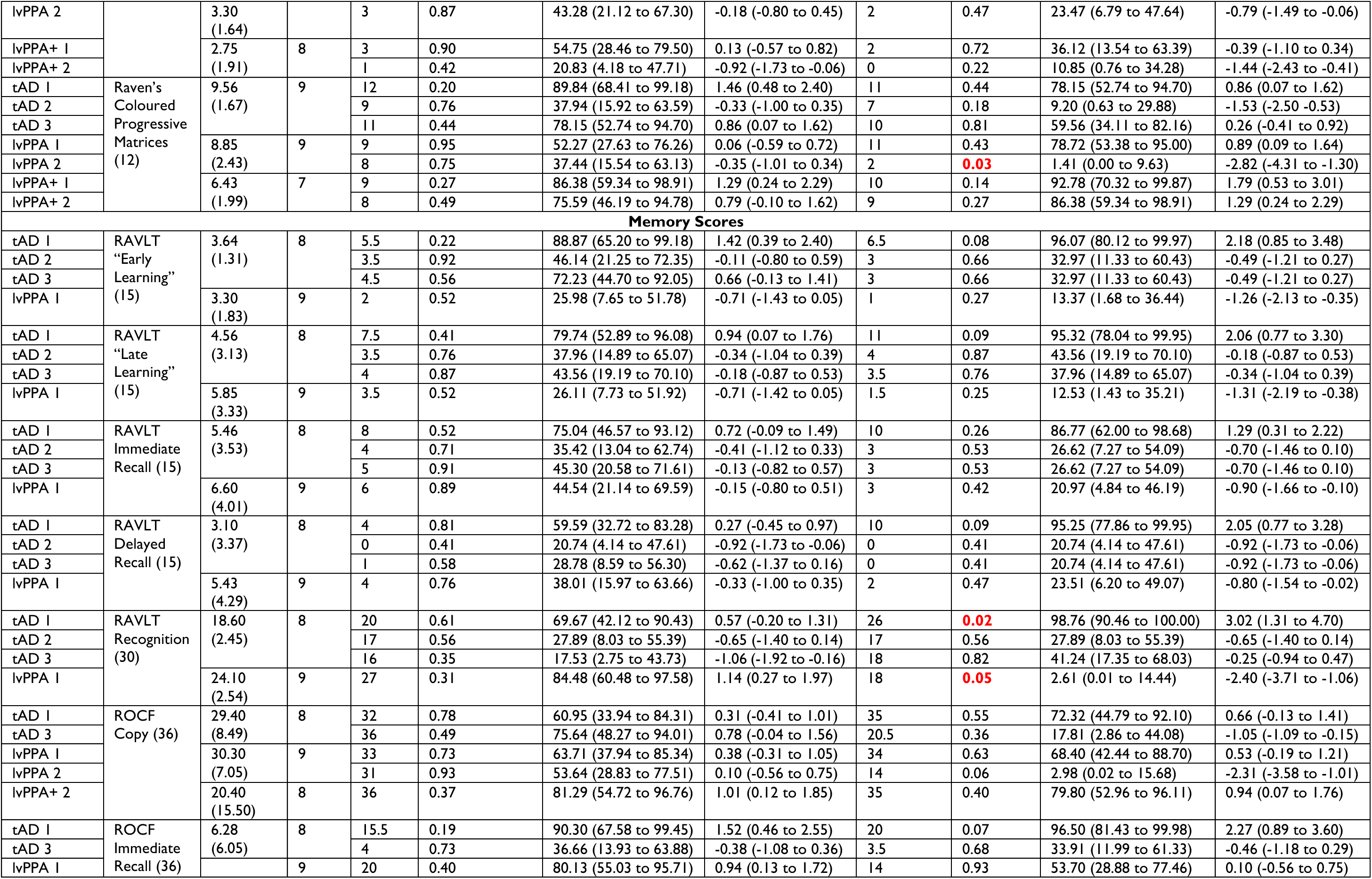
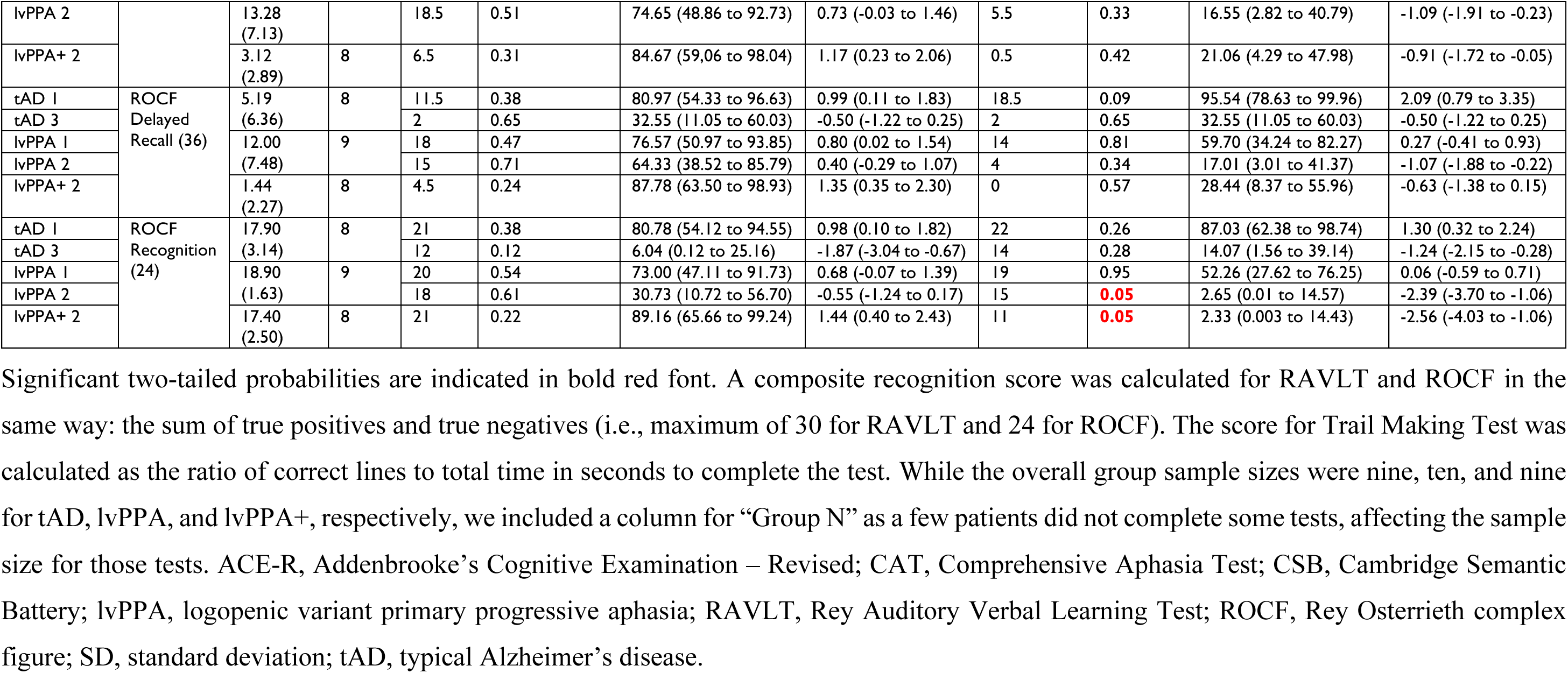
Bayesian point and interval estimates of effect sizes for each longitudinal case to group sample.

**Supplementary Fig. 1.**
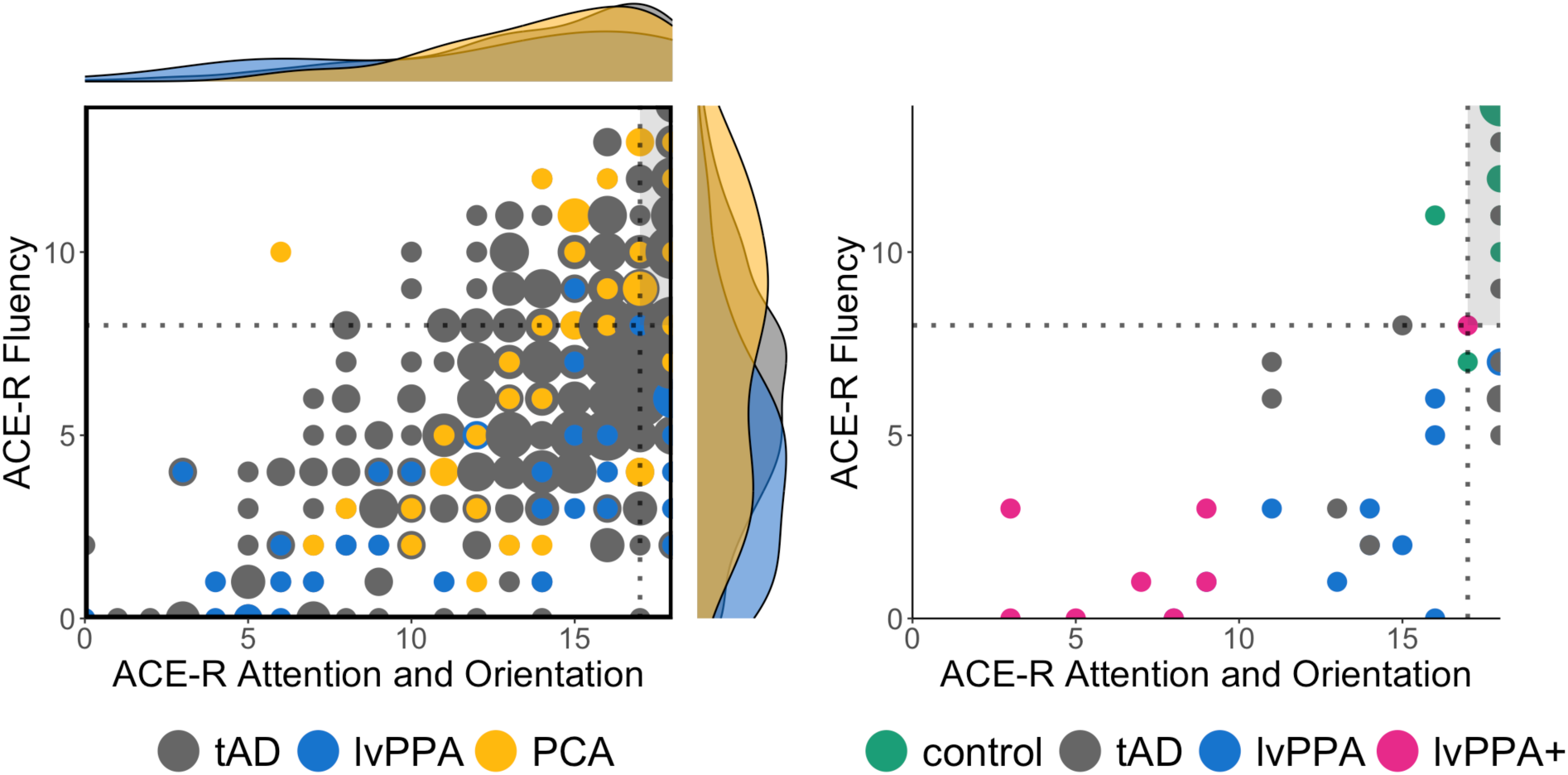
Scatterplots of ACE-R fluency and attention/orientation domains over the study population (left: control *n* = 12, tAD *n* = 9, lvPPA *n* = 10, lvPPA+ *n* = 8) and research participants who were classified as having an AD clinical phenotype from the memory clinics of the Cambridge University Hospital (right: AD *n* = 329, lvPPA *n* = 44, PCA *n* = 40). The dotted lines represent the ACE-R cutoff scores from published healthy control normative data ^28^ and the shaded grey sections portray the quadrant of normality. In the scatterplot on the right, the larger dots portray more participants who had the same scores and the accompanying raincloud plots on the sides show the density curves of the data distribution per group.

**Supplementary Fig. 2.**
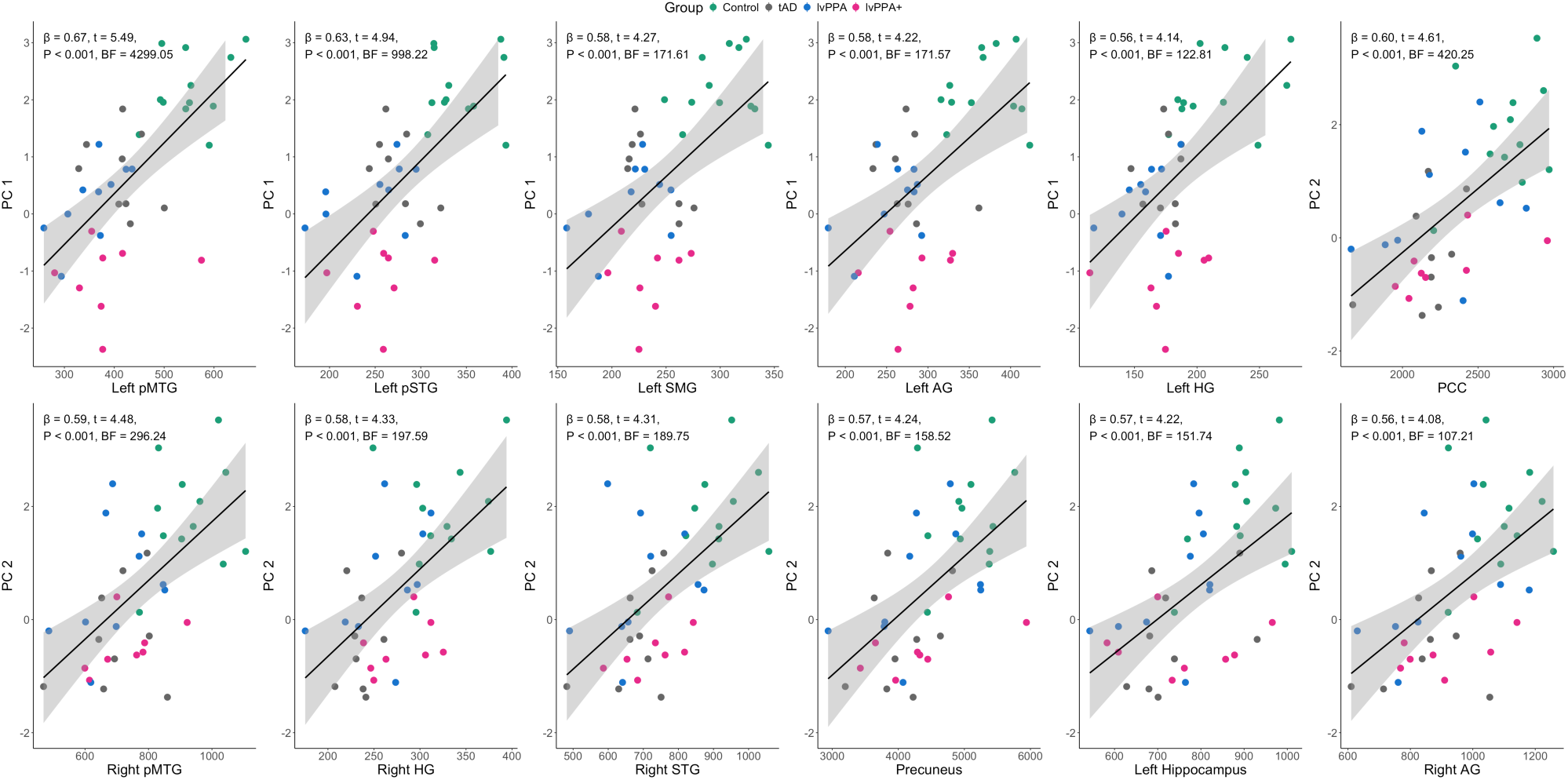
Associations between principal component (PC) scores and region of interest (ROI) grey matter volumes across the whole group that showed extreme Bayesian evidence. Only results showing extreme Bayesian evidence (BF > 100) are shown. AG, angular gyrus; BF, Bayes factor; HG, Heschl’s gyrus; PCC, posterior cingulate cortex; pMTG, posterior middle temporal gyrus; pSTG, posterior superior temporal gyrus; SMG, supramarginal gyrus.

**Supplementary Fig. 3.**
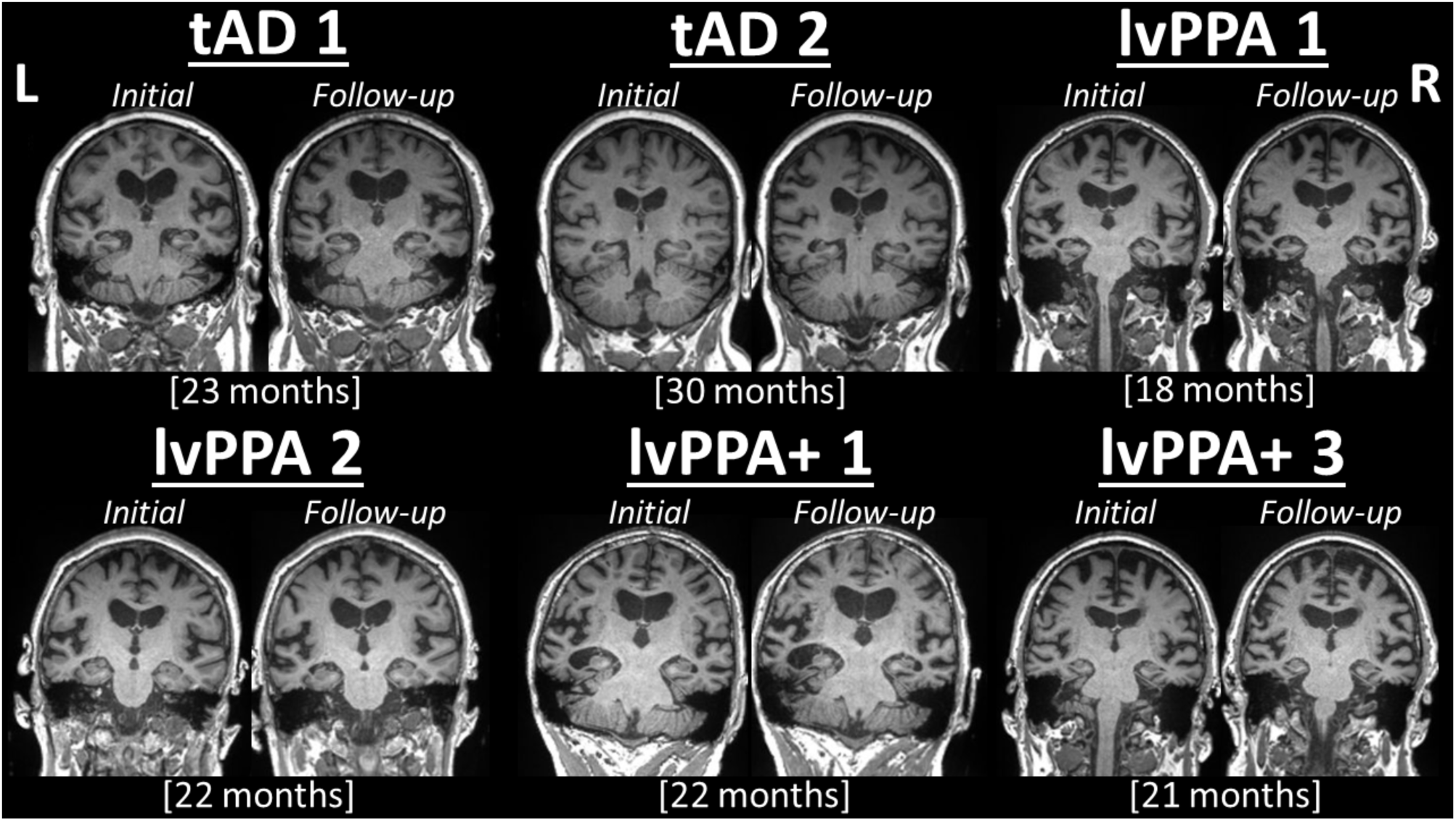
Neuroimaging for the longitudinal patients. Coronal MRI slices at initial and follow-up MRI scans are shown. Time since initial scan is indicated as months in square brackets.

